# Effectiveness of a novel non-intrusive continuous-use air decontamination technology to reduce microbial contamination in clinical settings: A multi-centric study

**DOI:** 10.1101/2021.08.25.21262596

**Authors:** Savitha Nagaraj, Sindhulina Chandrasingh, Sanju Jose, Sofia B, Sriram Sampath, Bhuvana Krishna, Indira Menon, Debosmita Kundu, Sandeepan Parekh, Deepak Madival, Vrinda Nandi, Arindam Ghatak

## Abstract

**Background:** Despite rigorous disinfection, fumigation and air treatment, infectious microbial load has been found to circulate and survive for significant durations in health care settings. This raises significant concerns for hospital acquired infections. We have developed a novel, hybrid, trap-and-kill airborne-microbicidal technology called “ZeBox” which is efficient in clearing 99.999% of airborne microbial load under controlled lab conditions. In this study we evaluate the clinical performance of the ZeBox in reducing airborne and surface microbial load in two independent hospital settings.

**Methods:** The studies were conducted in single bed and multi bed ICU of two hospitals. Airborne and surface microbial loads were collected at pre-determined sampling sites pre- and post-deployment of the ZeBox enabled device. The Normality of data distribution was determined using the Shapiro-Wilk test. Statistical significance was determined using Students’ T test and Mann-Whitney’s U test. Pathogenic and opportunistic organisms were characterized using 16S rDNA sequencing. Furthermore, the antibiotic sensitivity of the isolated organisms was tested against current treatments of choice across major antibiotic classes.

**Results:** Post-deployment, we found statistically significant reductions in both airborne and surface microbial load within the operating range of the ZeBox enabled technology. Across the both hospital ICUs, there was 90% reduction of airborne microbial load on average, and 75% reduction of surface microbial load on average, providing a low bioburden zone of roughly 10-15 feet diameter around the unit. These reduced microbial levels were maintained during the entire duration of device operation over several weeks. Many of the clinical isolates recovered from one of the hospitals were drug resistant, which highlighted the potential ability of ZeBox to eliminate drug-resistant microbes and thereby reduce the frequency of hospital acquired infections.

**Conclusions:** ZeBox enabled technology can significantly reduce a broad spectrum of microbial burden in air and on surfaces in clinical settings. It can thereby serve an unmet need in reducing the incidence of hospital acquired infections.

## Background

Human exposure to environmental organisms is associated with infectious disease transmission as well as allergic and non-allergic respiratory illnesses [1]. The most recent airborne Covid-19 pandemic highlights the continuous crisis caused by biological agents on public health. Even before the current pandemic, Healthcare-associated infections (HAIs) were a growing concern for clinical practice worldwide. Close to 2 million patients contract HAIs in the U.S. every year, out of which nearly 100,000 patients die [2]. The overall HAI rate is about three-fold higher in the developing world, with the risk of contracting device associated infections being as much as 15-19-fold higher [3]. These infections are often due to multidrug-resistant (MDR) bacteria, which have increasingly few treatment options [3, 4]. Additionally, the risk of contracting HAIs increases if the prior bed or room occupant suffered from infection; this risk is as much as four-fold higher for an Acinetobacter infection [5].

Microbial load in indoor environments can have various origins including shedding from occupant’s respiratory tract or skin, aerosolization from showers, or similar systems [6–8]. Quite often, bacteria, viruses, fungi, or fungal spores can also be resuspended from floors and deposited on interior surfaces distant from the source of contamination via airborne dispersion and can be further dispersed via contact with healthcare workers and through cross-contamination [9, 10]. Pathogenic microorganisms can survive in indoor environments for long periods of time [11] depending on the temperature and humidity, despite regular cleaning protocols instituted by healthcare spaces. The survivability of an organism is strongly dependent on the nature of the surface. For example, SARs-CoV-2 can survive between 2 hours to 28 days depending on the surface, ambient temperature, humidity and exposure to sunshine [12]. Extended contamination of surfaces can lead to the cumulative build-up of pathogens over time, particularly those that are resistant to surface or terminal room disinfection and can pose a significant hazard to the next patient [13]. Under typical heating, ventilation and air conditioning (HVAC) found in hospitals, Clostridium difficile spores, Vancomycin resistant Enterococcus (VRE), Methicillin resistant *Staphylococcus aureus* (MRSA) and *Acinetobacter baumannii* have been recovered after 4-5 months with surface contamination levels exceeding the number of bacteria or virions necessary for the transmission of infection [14, 15]. Causing even more concern for nosocomial spread, it has been found that Pseudomonas can linger on surfaces for as long as 16 months [14].

Hospitals, dental clinics, nursing homes and long-term care facilities typically see a large burden of pathogenic organisms posing a health risk to all occupants. Microbial contamination in hospital wards is concentrated in hard-to-reach surfaces such as the floor under beds and bed wheels as compared to higher levels of a room. This correlates both with the source of infection (patients in beds) and the fact that air trapped under beds and instruments is not efficiently cycled through wall mounted air purification units. There is a pressing need to design microbial decontamination devices that function near microbial reservoirs.

In dental clinics, aerosols generated through drills and scalers can potentially splatter or aerosolise and move within the indoor environment. Body fluids or blood from patients may harbour viruses (such as mumps, measles, rubella, HSV 1 and 2, HIV, HBV, SARS-CoV, influenza A H5N1, influenza A H1N1, MERS-CoV or SARs-CoV-2), or bacterial pathogens (such as *Mycobacterium tuberculosis* or *Legionella pneumophilla*) some of which can be transmitted through aerosols and water mists [16–22]. Studies in nursing homeand long-term care-home residents have shown that infections account for 27% to 63% of hospitalizations in the United States [23].

Reducing the load of pathogenic organisms to below infectious level is thus crucial to mitigate risk of infection, particularly in indoor spaces. The CDC recommends eliminating microbes at the source as they are produced as the first line of defense against the spread of infections [18]. This aspect has come into greater focus more recently with the rapid spread of coronavirus disease across the globe. Indoor air decontamination is an urgent medical need to maintain health and hygiene needs of occupants.

Currently available technologies for decontaminating room air belong to two broad categories: those which merely trap suspended matter in air (inanimate dust particles along with microbes) and those which are microbicidal. Each of these technologies have their merits and demerits, which have been reviewed in considerable detail by others [24–29].

We believe that an ideal air decontamination technology must trap and then kill microbes in situ, thus preventing any future growth and dissemination. While trap-and-kill microbicidal technologies are already available–UV irradiated filters, filters made of microbicidal fibers, and filters combined with plasma technology –they suffer from major demerits regarding flow permeability (which determines power consumption) and generation of toxic by-products during operation. We have developed a novel, hybrid, trap- and-kill airborne-microbicidal technology called “ZeBox”, which exploits the fact that microbes naturally possess net electric charge (characterized by their zeta-potential) and therefore they can be readily manipulated using an electric field. In ZeBox technology, a non-ionizing electric field is applied between electrode-plates on which unique microbicidal substrates are layered. The electric field plays two roles: it attracts the microbes to the microbicidal substrate and potentiates the substrate to instantaneously killing the trapped microbes [30].

In an enclosed test chamber under challenge conditions, ZeBox powered devices achieved 6-9-log_10_ reduction of a broad spectrum of microorganisms (airborne gram positive and gram-negative organisms of ESKAPE group, viruses, vegetative fungi and spores) in 10 minutes, a performance that is at least 1000-fold superior to that reported in the literature. In applications, which almost always consist of a space (enclosed or otherwise) with an unceasing flux of people and patients, a continuous and rapid-action microbicidal device is highly desirable. This is why the superior killing rate of ZeBox technology makes it unique for continuous real-time applications. In this paper, we evaluate the clinical performance of a ZeBox technology powered air decontamination device variant in reducing bacterial and fungal load in air and on surfaces in two independent hospital settings. We also delineate the typical pathogenic and opportunistic organisms found in these settings, to characterize the risk of nosocomial transmission to patients and health care staff.

## Methods

### A. Experimental Design for Testing Device Efficiency in Hospital ICUs

#### a) Single Bed and Multi Bed ICU Set Up

The studies were conducted in a single bed ICU and multi bed ICU located in two independent hospitals after approval from their Hospital Internal Ethics Committee. Both rooms were mechanically ventilated with filtered and tempered air at 22.6±1.9°C with no humidification. Housekeeping and nursing staff shared routine cleaning duties. Near-patient sites were cleaned by nurses twice daily at 7 am and 7 pm using wipes (Vernacare Tuffie™ wipes) and detergent (Hospec™). Terminal cleaning of the bed-space was performed following discharge. Samples were collected at specific locations identified as sampling sites between 2-3 pm, 7-8 hours post-cleaning of near-patient surfaces three or four times a week. In the Single Bed ICU, samples were collected four times a week over 11 weeks for determining baseline levels of contamination. The ZeBox powered air decontamination device was deployed at the end of the 11th week and samples were collected as before for another 10 weeks.

In the Multi bed ICU, samples were collected three times a week, over 13 weeks for determining baseline levels of contamination. The ZeBox powered air decontamination device was deployed at the end of the 13th week and samples were collected as before for another 13 weeks.

#### b) Selection of Sampling Sites

##### i. Single Bed ICU

The room had a dimension of 15 x 10 feet (Fig 1a). Indoor air samples were collected at positions S1 (medicine and reporting table) which was 10 feet away and S2 which was 6 feet away from the air decontamination device. Surface samples were collected from position S1 (medicine and reporting table) which was 10 feet away and position S3 (patient bed rails) which was 4 feet away from the ZeBox technology powered air decontamination device.

**Figure 01.**
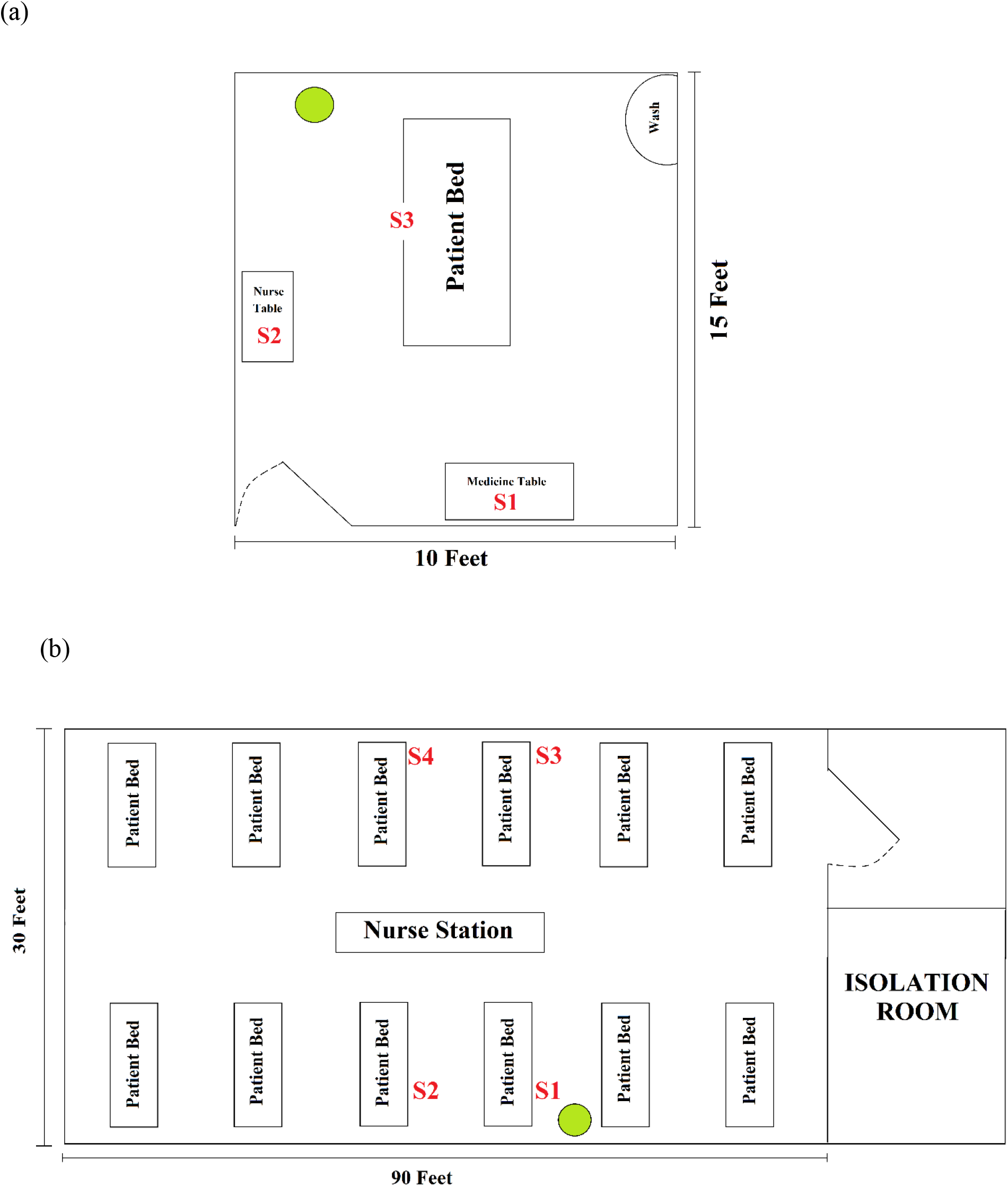
(a) ICU Room schematics for collecting air samples. The room has a dimension of 15×10 ft, samples were collected from positions S1 and S2 for quantification of total bacterial and fungal population. Position S1 and S2 were 10 Feet and 6 Feet away from the deployed device respectively. Sample positions S1 and S3 were 10 feet and 4 feet respectively from the deployed device and used for collecting surface microbial samples. The Green Circle denotes the position of the air decontamination unit. (b) The High Intensity Care Unit (HICU) room layout and the sampling positions (1,2,3,4). The HICU room has a dimension of 90ft x 30 ft, 12 patients can be treated in the HICU at any given point. The Green Circle denotes the position of the air decontamination unit. Position 1 was 2 feet away, Position 2 was 8 feet away, Position 3was 24 feet away and Position 4 was 26 feet away from the decontamination device. The deployed unit could effectively serve an area of 150sq. ft.

##### ii. Multi bed ICU

The room had a dimension of 30 feet x 90 feet. The sampling sites were chosen in consultation with the ICU staff to ensure the deployed device did not hinder movement and activities within the ICU. Sampling sites were selected such that two sites (positions 1 and 2) were proximal to the ZeBox technology powered air decontamination unit and served as sites on which the direct effect of the device could be monitored. Two other sites (positions 3 and 4) were distal to the ZeBox technology powered air decontamination uni and served as the control sampling sites (Figure 1b). The deployed unit (marked by a green circle) could effectively serve an area of 150sq. ft.

### B. Sampling of air for viable microbes

A handheld air sampler (SAS Super ISO 100, VWR), which could sample 100 liters of air per minute, was used to collect air samples. A fixed volume of air was sampled using the air-sampler. Tryptic Soy Agar and Sabouraud dextrose agar plates were used to sample bacteria and fungi, respectively, from the air. Plates were placed in and removed from the air-sampler in an aseptic manner. Plates were incubated at 25±2^0^ C (for fungal cultivation) and 37±2^0^ C (for bacterial cultivation) for 48 hours. After incubation, the number of colonies were enumerated and converted to CFU/m^3^ using statistical conversion provided by the manufacturer. Control plates were used to ensure the sterility of the entire process.

### C. Surface sampling

A cotton swab was moistened with sterile phosphate-buffered saline (1X PBS; pH 7.2) solution using aseptic technique to prevent cross-contamination and was used to wipe a surface of 100cm^2^ area as mentioned in CDC Guideline (EMERGENCY RESPONSE RESOURCES https://www.cdc.gov/niosh/topics/emres/unp-envsamp.html). The sampled swab was placed in a sterile conical vial containing 1ml of sterile phosphate-buffered saline (PBS) solution. The entire 1ml solution was then plated on to Tryptic Soy Agar and Sabouraud dextrose agar plates for quantification of bacteria and fungi, respectively. Plates were incubated at 25±2^0^ C (for fungal cultivation) and 37±2^0^C (for bacterial cultivation) for 48 hours. Post-incubation, the number of colonies that appeared were enumerated. Control plates were used to ensure the sterility of the entire process.

### D. Microbial identification from Multi Bed Hospital ICU using 16S rDNA sequencing

Bacterial population from air were collected using a handheld air-sampler on a TSA plate and incubated for 48 hours, allowing the collected microbes to grow and form visible colonies. Colonies were first screened based on their morphological characteristics, (viz texture, color, shape, and elevation) and grouped accordingly. Individual isolates from these groups were then picked for 16S rDNA sequencing. The 16S rRNA gene region of bacterial genomic DNA was amplified using universal bacterial primers. Each of the PCR reaction systems contained 2 *μ*l of Forward primer (0.4 *μ*M), 5’-AGR GTT TGA TCM TGG CTC AG-3’, 2 *μ*l of Reverse primer (0.4 *μ*M), 5’-GGY TAC CTT GTT ACG ACT T-3’, 5 *μ*l of PCR Green buffer (1X), 1.5 *μ*l of MgCl_2_ (1.5 mM), 2.5 *μ*l of dNTP (0.1 mM), 0.2 *μ*l of Taq DNA Polymerase (1 unit), and approximately 7.5 ng of DNA template. It was followed by the addition of 20 *μ*l of mineral oil on top of each PCR reaction mixture. The *E. coli* ATTC 25922 DNA was used as the positive control and PCR master mix with Milli-Q water was used as a negative control. PCR amplicons were sequenced at the The Bangalore Biocluster Next Generation Genomics Facility (TIFR-NCBS, Bangalore, India). The sequence trace files were assembled, trimmed, aligned and manually checked using Bionumerics software 6.0 (Applied. Maths. Sint-Martens Latem, Belgium), and the sequences were classified using the Classifier and SeqMath tools [31] at the Ribosomal Database Project (RDP) and BLASTn databases via the online interface at National Center of Biotechnology and Information (NCBI). Sequenced genes were aligned using Clustal Omega (https://www.ebi.ac.uk/Tools/msa/clustalo/) and taxonomical analysis were carried out simultaneously.

### E. Antibiotic sensitivity test of microbes collected from Multi Bed HICU

Single colony of each strain was grown in M9 medium. All test compound stocks and dilutions were prepared in DMSO. Serial two-fold dilutions of antibiotics were prepared separately, with concentrations ranging from 2 mg/mL to 0.015 mg/mL. To 150 µl (3– 7×10^5^CFU/ml) of bacterial culture in 96 well microtiter plates, 3 μL compound from each of the dilutions was added into respective wells to obtain final concentrations ranging from 40 µg/mL to 0.3 µg/mL of the test compounds. Media control, culture control and appropriate reference drug controls were included. The plates were packed in gas permeable polythene bags and incubated at 37 °C overnight. Growth was monitored by checking absorbance at 600nm (A600). Minimum inhibitory concentration (MIC) was taken as the concentration that resulted in a growth inhibition of ≥80%.

### F. Statistical Analysis of Data

All data sets were tested for normal distribution using Shapiro-Wilk test (SW test), following which a non-parametric test, the “Mann-Whitney’s U test” (MWU test) was conducted. The details are available in the Supplementary material.

## Results

### A. Validation in Single Bed ICU

#### a) Airborne microorganisms

The environmental microbial load was monitored in a single bed ICU room when occupied by patients. Air samples for monitoring baseline load were collected over a period of 11 weeks to enumerate microbial distribution at two locations within the room. This was followed by ZeBox technology powered air decontamination unit deployment and sample collection over a subsequent period of 10 weeks with the first sample collection within a period of 3 hours after device deployment.

The airborne bacterial load before deployment was more or less similar at both the medicine table and nurse station, and showed roughly a four-fold intra-day variation over the period of 11 weeks (Figure 2, Table 1), ranging from 580-3000 CFU/m^3^ (average 1168 CFU/m3) at the medicine table (S1) and 80-1910 CFU/m^3^ (average 1147 CFU/m3) at the nurse station (S2). Similarly, the airborne fungal load before deployment showed roughly a three-fold variation day to day over 11 weeks, but on some days, the CFU counts were as high as four to seven-fold from the average daily counts (Figure 3, Table 1). Airborne fungal counts at the medicine table (S1) ranged from 78-688 CFU/m^3^ (average 157 CFU/m^3^) and 72-698 CFU/m^3^ (average 168 CFU/m^3^) at the nurse station (S2) before device deployment.

**Figure 2:**
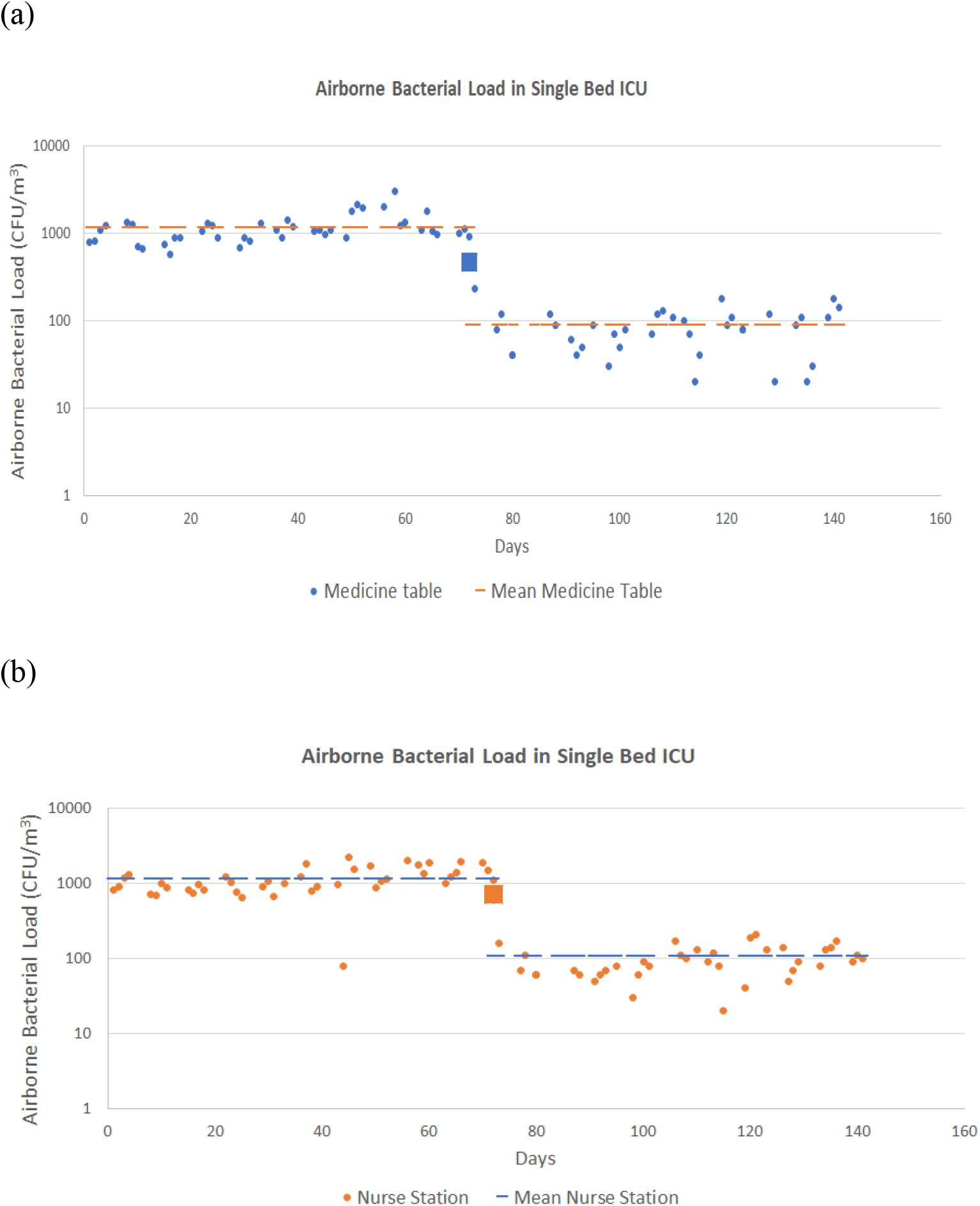
Airborne Bacterial loads at two positions in the Single Bed ICU. (a) Position S1, Medicine table and (b) position S2, Nurse Station. The average load before and after ZeBox technology powered air decontamination unit deployment is depicted by a line for both positions. The ZeBox powered air decontamination unit was deployed on day 72 and the first sample was taken within 3 hours of deployment. The microbial count for that time point is depicted by the square.

**Figure 3:**
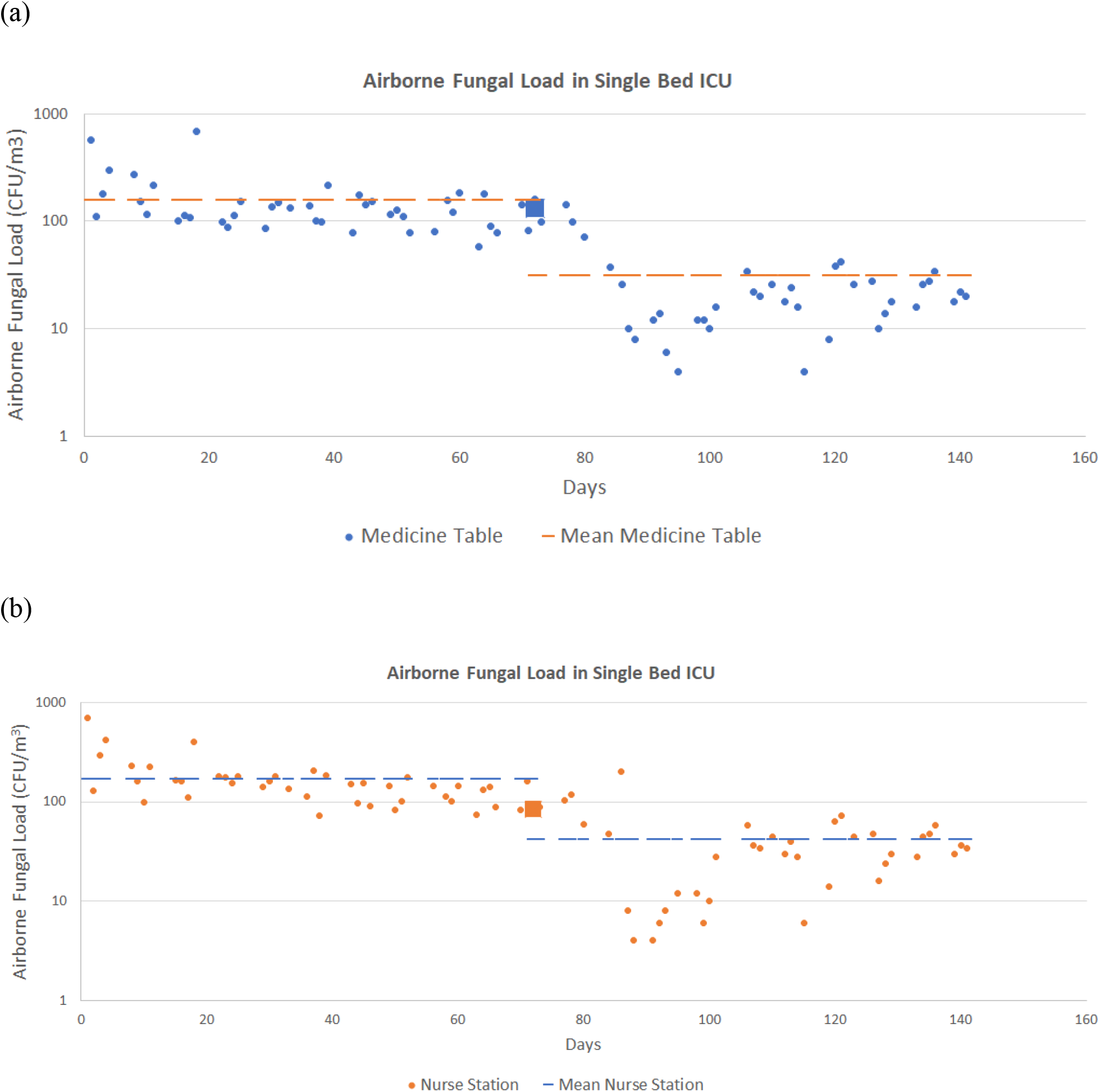
Airborne Fungal Loads at two positions in the Single Bed ICU. (a) Position S1, Medicine table and (b) position S2, Nurse Station. The average load before and after ZeBox technology powered air decontamination unit deployment is depicted by a line for both positions. The ZeBox powered air decontamination unit was deployed on day 72 and the first sample was taken within 3 hours of deployment. The microbial count for that time point is depicted by the square.

**Table 1:** Descriptive statistics for airborne and surface microbial load in a single bed ICU. The airborne bacterial and fungal loads were measured at medicine table and nurse station. The surface bacterial and fungal loads were measured at medicine table and bed rails.

|  |  | Airborne Bacterial load<br>(CFU/m<sup>3</sup>) |  | Airborne Fungal Load<br>(CFU/m<sup>3</sup>) |  |
| --- | --- | --- | --- | --- | --- |
|  |  | Medicine table | Nurse Station | Medicine table | Nurse Station |
| Pre-Deployment | Range | 580-3000 | 80-1910 | 78-688 | 72-698 |
|  | Mean | 1168 | 1147 | 157 | 168 |
|  | Median | 1080 | 1020 | 128 | 146 |
| Post Deployment | Range | 0-470 | 0-710 | 4-144 | 4-202 |
|  | Mean | 89 | 108 | 31 | 42 |
|  | Median | 80 | 90 | 20 | 34 |
|  | % Reduction | 92 | 91 | 80 | 75 |

|  |  | Surface Bacterial Load<br>(CFU/m<sup>2</sup>) |  | Surface Fungal Load<br>(CFU/m<sup>2</sup>) |  |
| --- | --- | --- | --- | --- | --- |
|  |  | Medicine Table | Bed rail | Medicine Table | Bed rail |
| Pre-Deployment | Range | 1500-9400 | 200-10500 | 400-1100 | 0-1300 |
|  | Mean | 4681 | 1667 | 713 | 581 |
|  | Median | 3950 | 750 | 700 | 550 |
| Post Deployment | Range | 800-1900 | 200-1900 | 0-300 | 0-300 |
|  | Mean | 1269 | 913 | 113 | 150 |
|  | Median | 1100 | 700 | 100 | 200 |
|  | % Reduction | 73 | 45 | 84 | 74 |

After deployment of the decontamination device, the airborne bacterial load was reduced to 0-470 CFU/m^3^ at the medicine table (S1) (average 89 CFU/m^3^), and 0-710 CFU/m^3^ at the nurse station (average 108 CFU/m^3^) (Figure 2, Table 1). This accounts for a 92% reduction of airborne bacterial load at the medicine table (S1) and a 91% reduction at the nurse station (S2). The fungal load after deployment reduced to 4-144 CFU/m^3^ (average 31 CFU/m3) at the medicine table and 4-202 CFU/m^3^ (average 42 CFU/m^3^) at the nurse station (Figure 3, Table 1). This accounts for 80% reduction of fungal load at the medicine table and a 75% reduction at the nurse station. Both airborne bacterial and fungal load dropped significantly within a period of 3 hours post deployment of the device. The device was in continuous operation for the remaining duration of the study.

#### b) Surface microorganisms

Surface samples were collected from the medicine storage table and patient bed rail in the single ICU bed cubicle. Bacterial load before deploying the decontamination device ranged from 1500-9400 CFU/m^2^ and 200-10500 CFU/m^2^ on medicine table (S1) and patient bed rails (S3) respectively (Supplementary Figure S1, Table 1). After device deployment, surface bacterial load reduced to 800-1900 and 200-1900 on medicine table and patient bed rails respectively. This accounts for a 73% reduction of surface bacterial load on the medicine table. However, there was considerable scatter in the bacterial load on the bed rails both before and after device deployment and no significant reduction of surface bacterial load (∼45% reduction of the mean surface load).

Surface Fungal load before deploying the decontamination device ranged from 400-1100 CFU/m^2^ and 0-1300 CFU/m^2^ on medicine table and patient bed rails, respectively. Post-deployment of the decontamination device, the surface fungal load was reduced to 0-300 CFU/m^2^ on both medicine table and patient bed rails (Supplementary Figure S2, Table 1). This accounts for 84% reduction of airborne bacterial load at the medicine table and a 74% reduction on the bed rails.

### B. Validation in a Multi Bed HICU

#### a) Airborne microorganisms

The environmental microbial load was monitored in a functional multi-bed HICU room occupied by patients with regular movement of hospital personnel. Air samples were collected as mentioned previously and total culturable microbial load was enumerated. Baseline samples were collected over a period of thirteen weeks to understand the microbial distribution at various positions in the room. This was followed by device deployment and sample collection over a period of a further thirteen weeks. Depending on the position sampled, the bacterial and fungal load in the air before deployment showed considerable variability over time. Airborne bacterial load before device deployment ranged from 58-398 CFU/m^3^ while the fungal load ranged from 14-130 CFU/m^3^ across the four positions (Table 2).

**Table 2:**
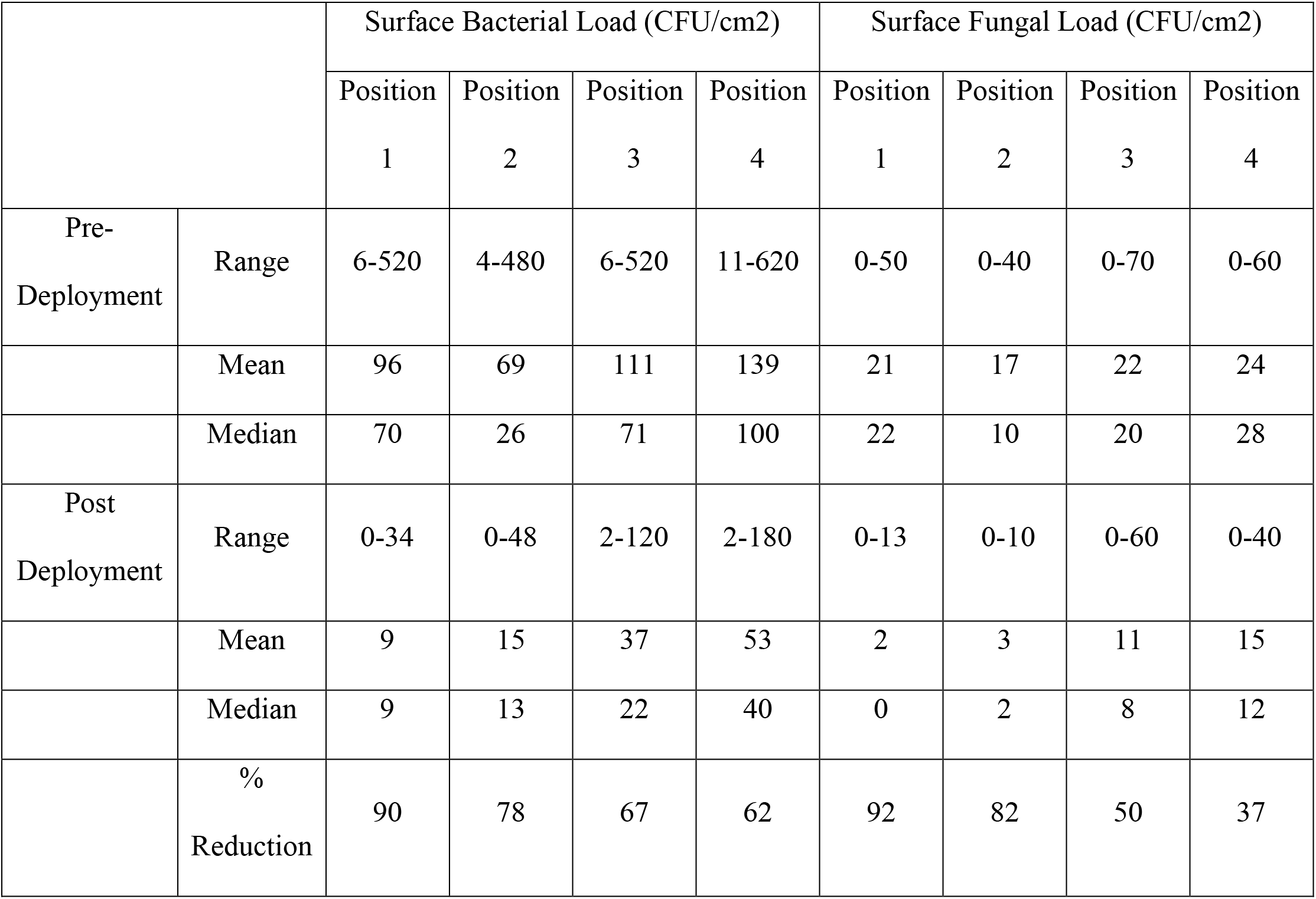
Descriptive statistics for airborne and surface microbial load in a multi bed ICU. The airborne bacterial and fungal loads were measured at four positions. Position 1 and 2 were 2 feet and 8 feet away respectively while positions 3 and 4 were 25 feet and 26 feet away respectively from the decontamination device.

After deployment of the decontamination device, the airborne microbial load was reduced to 0-210 CFU/m^3^ and 0-98 CFU/m^3^ for bacterial and fungal population, respectively, across the four sampling positions. The maximum reduction in bacterial load in air was shown at Position 1 (0-66 CFU/m^3^) and Position 2 (0-44 CFU/m^3^), which were 2 and 8 feet away from the ZeBox technology powered air decontamination unit, than at Positions 3 (20-210 CFU/m^3^) and Positions 4 (24-208 CFU/m^3^), which were 24 and 26 feet away from the device (Figures 4, Table 2). This accounted for a 96-97% reduction in airborne bacterial load at Positions 1 and 2, but only 40-44% reduction in airborne bacterial load at Positions 3 and 4.

**Figure 4:**
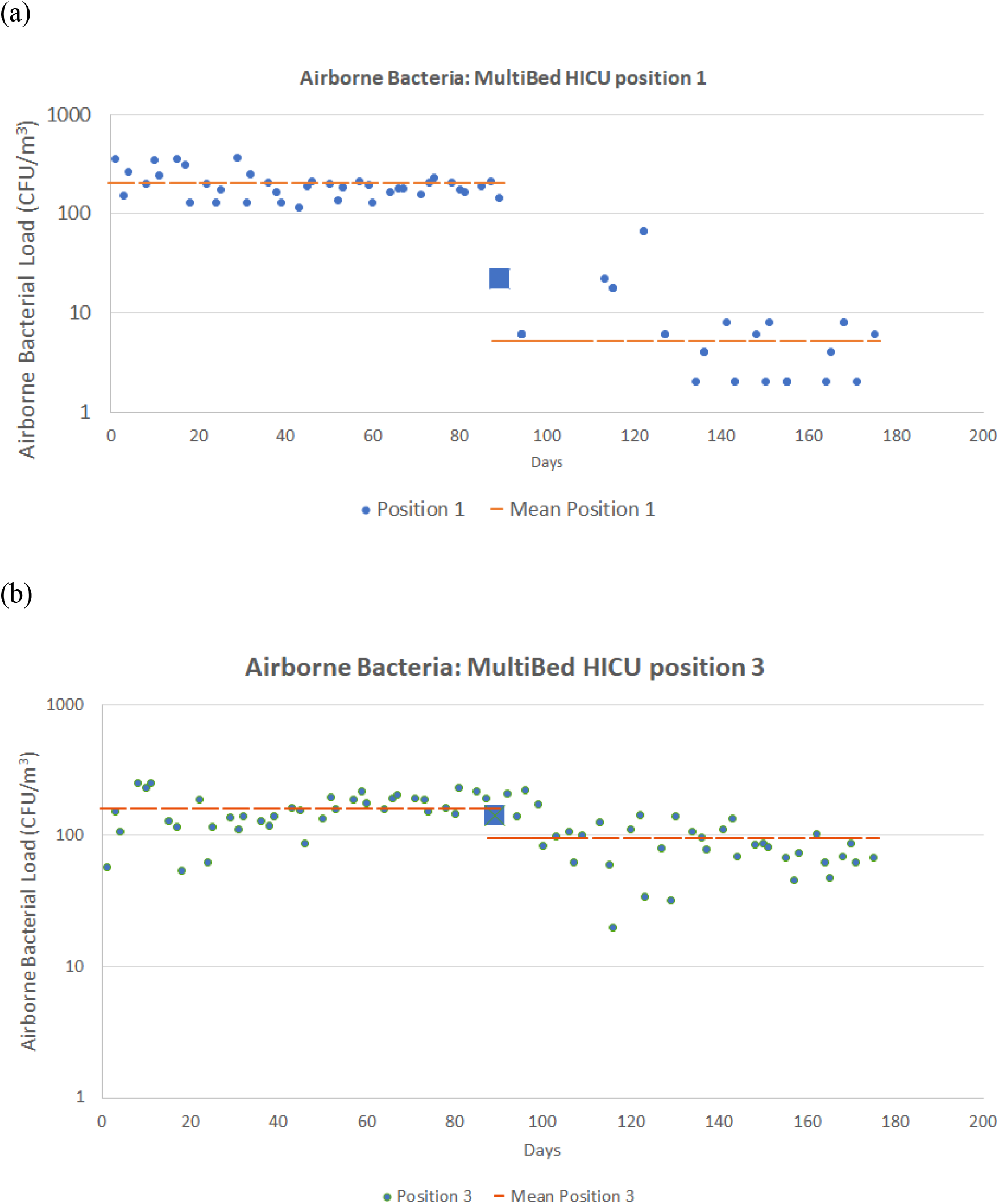
Airborne Bacterial load in Multi bed ICU. Active sampling was carried out from four positions. Positions 1 and 2 were 2 feet and 8 feet away, respectively, from the ZeBox technology powered air decontamination unit. Positions 3 and 4 were 25 feet and 26 feet away from the air decontamination unit. The ZeBox powered air decontamination unit was deployed on day 89 and the first sample was taken within 3 hours of deployment. The microbial count for that time point is depicted by a square. Graphs for positions 1 (Fig. 4a) and 3 (Fig. 4b) are shown here for comparison. Graphs for position 2 and 4 can be found in the supplementary material (Supplementary Figure 2A,2B).

The trend for airborne fungal load also showed a similar pattern. The maximum reduction was shown at Positions 1 and 2 (0-18 CFU/m^3^) as compared with Positions 3 (0-98 CFU/m^3^) and Positions 4 (0-58 CFU/m^3^). This accounted for 93-94% reduction of airborne fungi at Positions 1 and 2, but only 51-53% reduction at Positions 3 and 4 (Figure 5, Table 2).

**Figure 5:**
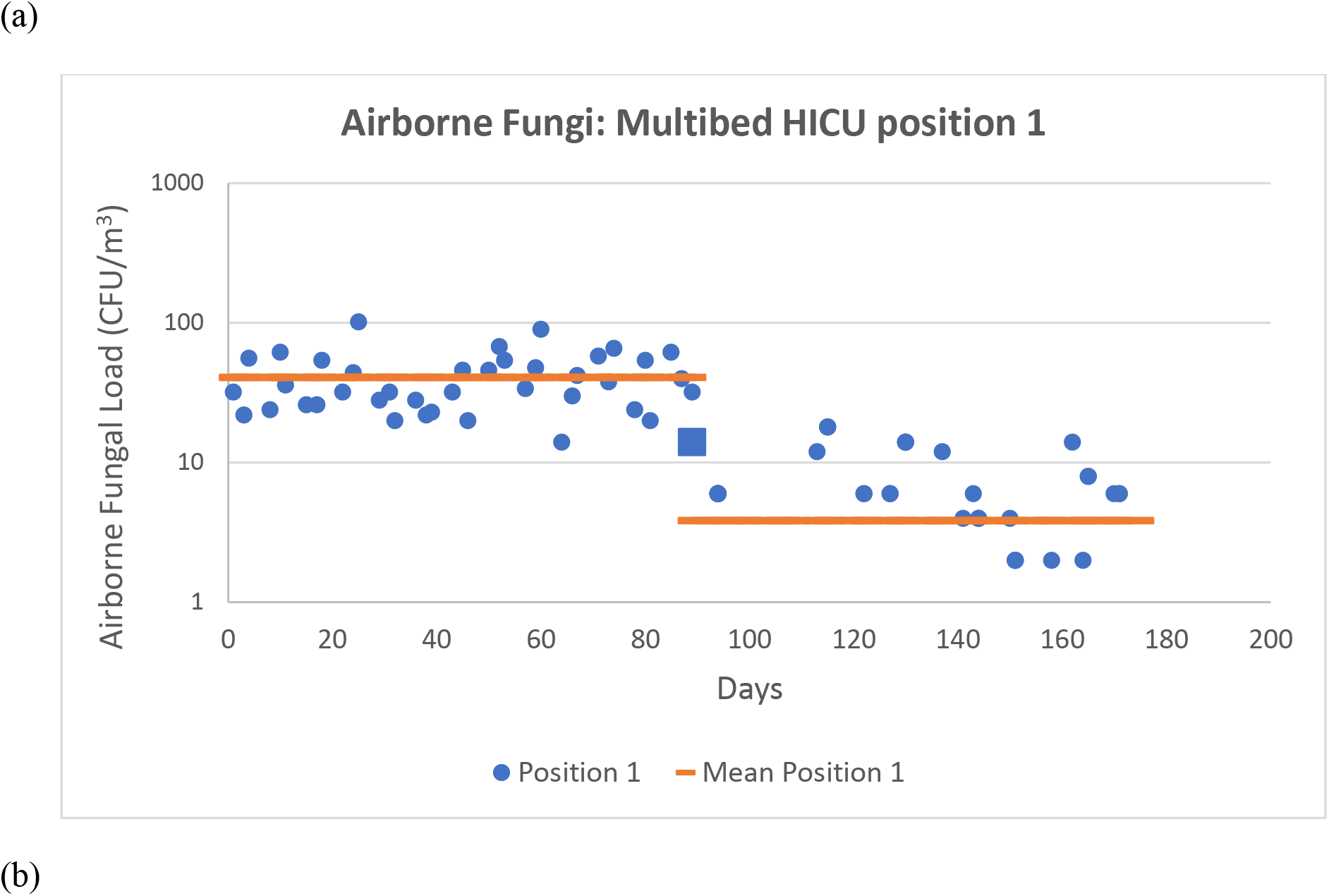
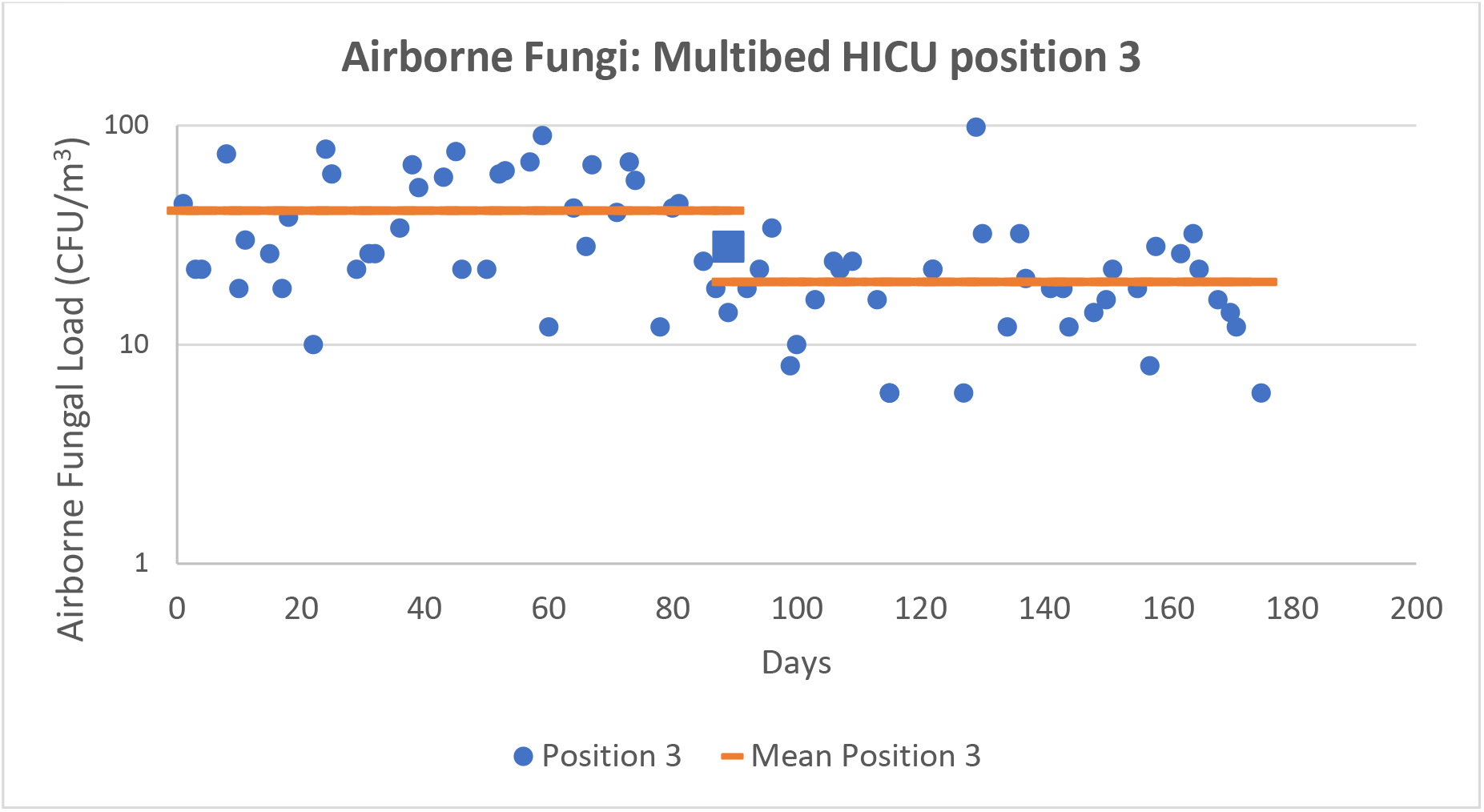
Assessment of Airborne fungal load in Multi bed ICU. Active sampling was carried out from four positions. Positions 1 and 2 were 2 feet and 8 feet away, respectively, from the ZeBox technology powered air decontamination unit. Positions 3 and 4 were 25 feet and 26 feet away from air decontamination unit. The ZeBox powered air decontamination unit was deployed on day 89 and the first sample was taken within 3 hours of deployment. The microbial count for that time point is depicted by a square. Graphs for positions 1 (Fig. 8 a) and 3 (Fig 8 b) are shown here for comparison. Graphs for position 2 and 4 can be found in the supplementary material.

**Figure 6:**
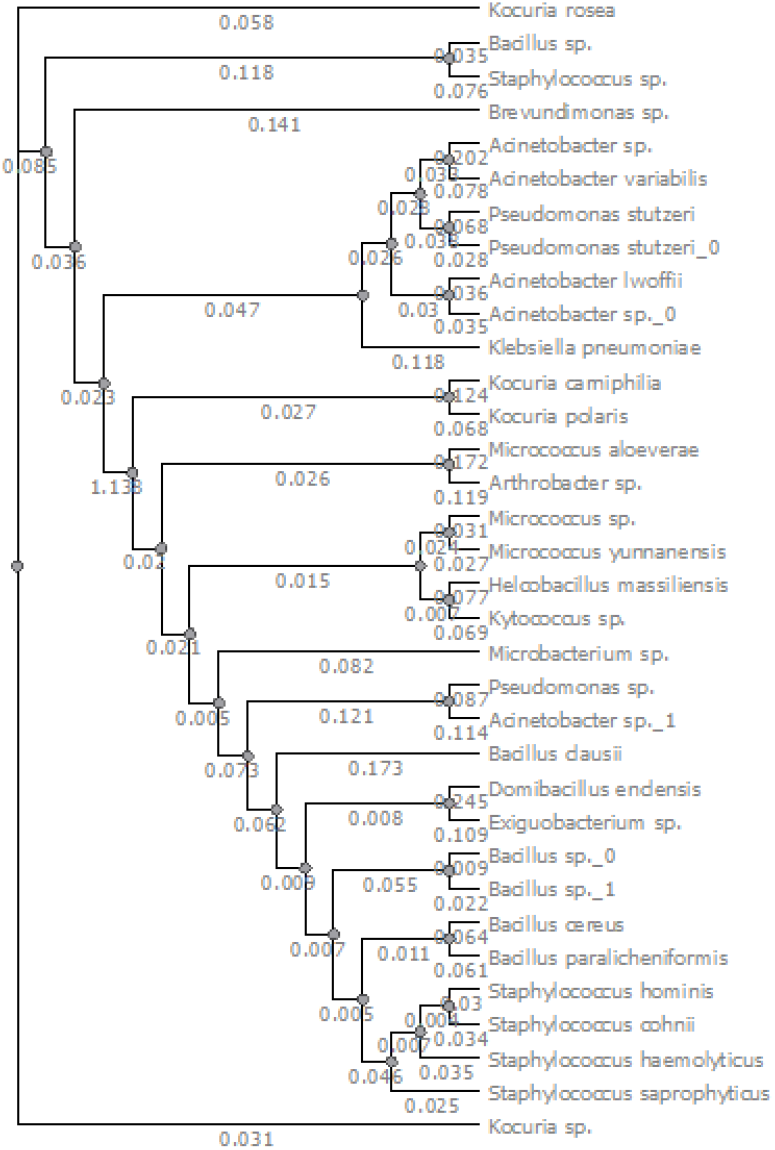
Phylogenetic tree of organisms isolated

#### a) Surface microorganisms

The surface microbial load was monitored using techniques mentioned previously. Samples were collected from four different positions in the HICU room. Patient bed rails were selected as sampling locations.

Surface bacterial load before device deployment ranged from 6-620 CFU/cm^2^ while the fungal load ranged from 0-70 CFU/cm^2^ across the four positions. After deployment of the decontamination device, the microbial load was reduced to 0-180 CFU/cm^2^ and 0-60 CFU/cm^2^ for bacterial and fungal population, respectively, across the four sampling positions. As before, the maximum reduction in surface bacterial load was shown at Position 1 (0-34 CFU/cm^2^) and Position 2 (0-48 CFU/cm^2^), which were 2 and 8 feet away from the device, while a lowered reduction was observed at Positions 3 (2-120 CFU/cm^2^) and Positions 4 (2-180 CFU/cm^2^), which were 24 and 26 feet away from the device. This accounted for a 78-90% reduction in surface bacterial load at Positions 1 and 2, but only 62-67% reduction in surface bacterial load at Positions 3 and 4. Similarly, the surface fungal load showed maximum reduction at Position 1 (0-13 CFU/cm^2^) and Position 2 (0-10 CFU/cm^2^) and lower reductions at Position 3 (0-60 CFU/cm^2^) and Position 4 (0-40 CFU/cm^2^). This accounted for 82-92% reduction of surface fungal load at Positions 1 and 2, and 37-50% reduction in surface fungal load at Positions 3 and 4 (Supplementary Figure 03, Table 02).

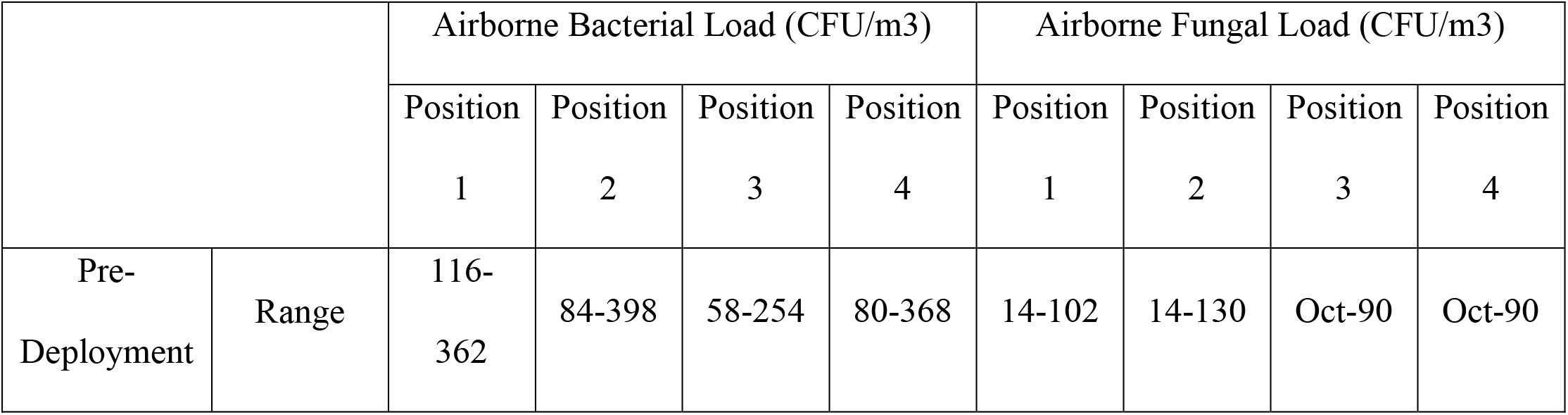
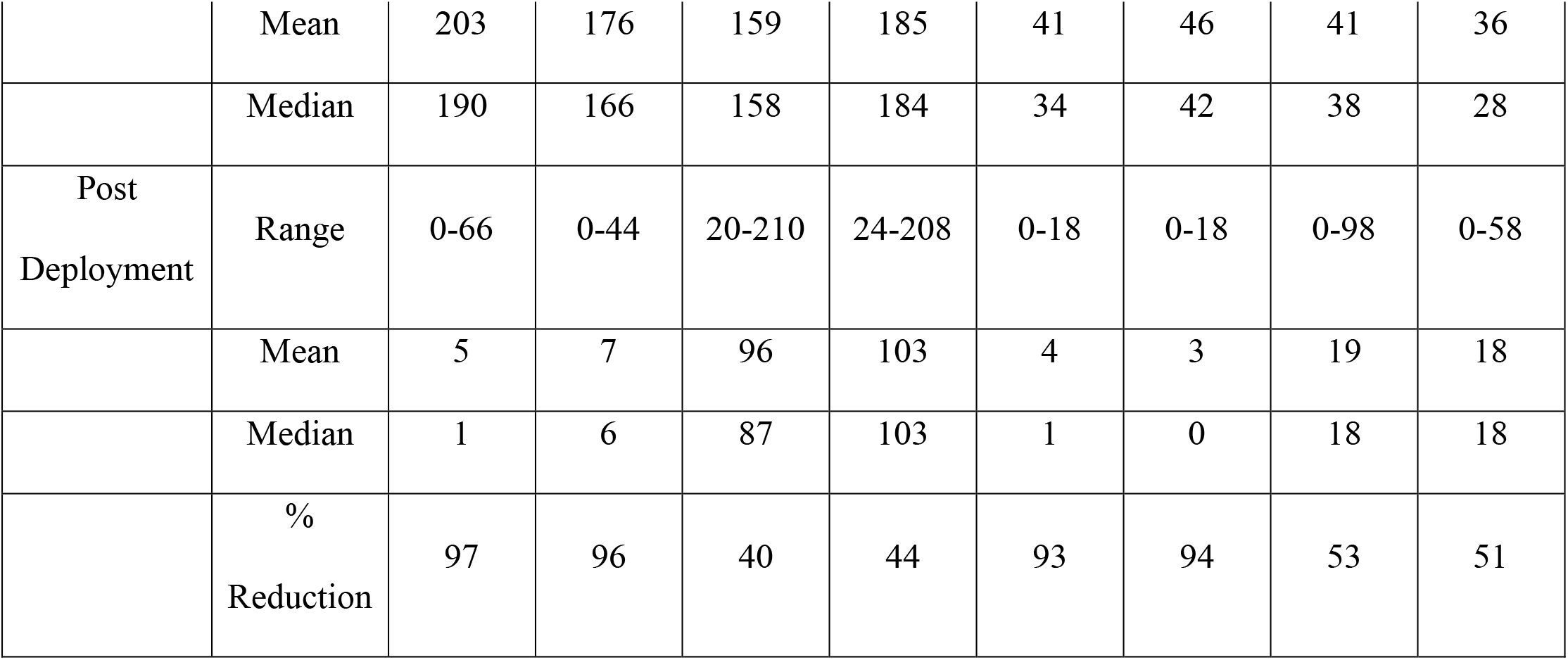

|  |  | Airborne Bacterial Load (CFU/m3) |  |  |  | Airborne Fungal Load (CFU/m3) |  |  |  |
| --- | --- | --- | --- | --- | --- | --- | --- | --- | --- |
|  |  | Position | Position | Position | Position | Position | Position | Position | Position |
|  |  | 1 | 2 | 3 | 4 | 1 | 2 | 3 | 4 |
| Pre-Deployment | Range | 116-362 | 84-398 | 58-254 | 80-368 | 14-102 | 14-130 | Oct-90 | Oct-90 |
|  | Mean | 203 | 176 | 159 | 185 | 41 | 46 | 41 | 36 |
|  | Median | 190 | 166 | 158 | 184 | 34 | 42 | 38 | 28 |
| Post<br>Deployment | Range | 0-66 | 0-44 | 20-210 | 24-208 | 0-18 | 0-18 | 0-98 | 0-58 |
|  | Mean | 5 | 7 | 96 | 103 | 4 | 3 | 19 | 18 |
|  | Median | 1 | 6 | 87 | 103 | 1 | 0 | 18 | 18 |
|  | %<br>Reduction | 97 | 96 | 40 | 44 | 93 | 94 | 53 | 51 |

### C. Microbial identification from Multi Bed Hospital ICU using 16S rDNA sequencing

Following sequencing, alignment and taxonomical analysis on the sequenced genes, the organisms were identified and classified as pathogenic and non-pathogenic. As shown in Table 3, the pathogenic organisms identified were *Bacillus cereus, Acinetobacter baumanii), Acinetobacter lwoffii (*potentially opportunistic pathogen), *Klebsiella pneumoniae, Brevundimonas sp. (*rare case of opportunistic pathogen), *Pseudomonas stutzeri* (an opportunistic pathogen), *Staphylococcus saprophyticus and Staphylococcus haemolyticus, hominis, cohnii* (emerging opportunistic pathogens). In addition, over half a dozen non-pathogenic organisms were identified.

**Table 3:** Antibiotic sensitivity of isolated organisms against seven antibiotics shown as MIC (µg/ml).

| Organism | Ciprofloxacin | Meropenem | Ceftazidime | Azithromycin | Rifampicin | Ampicillin | Linezolid |
| --- | --- | --- | --- | --- | --- | --- | --- |
|  | MIC (µg/ml) |  |  |  |  |  |  |
| **Pathogenic** |  |  |  |  |  |  |  |
| Acinetobacter sp. | 0.25 | 0.08 | 10 | 40 | 0.08 | 1.3 | 5 |
| Acinetobacter lwoffii | 0.25 | 0.08 | 10 | 40 | 0.08 | 1.25 | 5 |
| Bacillus cereus | 0.06 | 0.08 | 10 | 40 | 0.08 | 2.5 | 1.3 |
| Bacillus sp. | 0.06 | 0.08 | 10 | 40 | 0.08 | 2.5 | 1.3 |
| Klebsiella pneumoniae | 0.02 | 0.04 | 0.15 | 0.3 | 0.63 | 0.15 | 0.15 |
| Pseudomonas stutzeri | 0.02 | 0.04 | 0.15 | 0.3 | 0.63 | 0.15 | 0.2 |
| Pseudomonas sp. | 0.06 | 0.08 | 10 | 40 | 0.08 | 2.5 | 1.3 |
| Staphylococcus cohnii | 0.5 | 2.5 | 10 | 40 | 0.08 | 10 | 1.3 |
| Staphylococcus haemolyticus | 0.5 | 0.31 | 10 | 40 | <0.02 | 0.3 | 2.5 |
| Staphylococcus hominis | 0.5 | 0.31 | 10 | 40 | <0.02 | 0.3 | 2.5 |
| Staphylococcus saprophyticus | 0.25 | 0.08 | 10 | 40 | 0.08 | 1.25 | 5 |
| **Non-Pathogenic / Opportunistic** |  |  |  |  |  |  |  |
| Bacillus clausii | 0.06 | 0.08 | 10 | 40 | 0.08 | 2.5 | 1.3 |
| Bacillus paralicheniformis | 0.06 | 0.08 | 10 | 40 | 0.08 | 2.5 | 1.3 |
| Domibacillus enclensis | 0.06 | 0.08 | 10 | 40 | 0.08 | 2.5 | 1.3 |
| Helcobacillus massiliensis | 0.06 | 0.08 | 10 | 40 | 0.08 | 2.5 | 1.3 |
| Kocuria polaris | 0.02 | 0.04 | 0.15 | 0.3 | 0.63 | 0.15 | 0.15 |
| Kytococcus sp. | 4 | 10 | 10 | 40 | 40 | 40 | 2.5 |
| Micrococcus aloeverae | 4 | 10 | 10 | 40 | 40 | 40 | 2.5 |
| Micrococcus yunnanensis | 4 | 10 | 10 | 40 | 40 | 40 | 2.5 |
| Microbacterium sp. | 0.25 | 0.08 | 10 | 40 | 0.08 | 1.3 | 5 |

### D. Antibiotic sensitivity test of microbes collected from Multi Bed HICU

The Minimum inhibitory concentration (MIC) of each of seven antibiotics was tested against each isolated organism. The seven antibiotics chosen represent current treatment choices across the various classes of available antibiotics. The sensitivity of the organisms to these antibiotics is tabulated in Table 3. Among the clinical isolates characterized, several strains were resistant to Ceftazidine, Azithromycin and Ampicillin. The isolates were relatively more sensitive to meropenem and linezolid. Most isolates were highly sensitive to Ciprofloxacin and Rifampicin. The *Kytococcus* and *Micrococcus* isolates seemed resistant practically to all antibiotics tested and had a modest sensitivity towards Linezolid. *Brevundimonas* could not be cultured for susceptibility testing.

### G. Statistical Analysis of Data

Statistical analysis was carried out on the data set after leaving out the transition period, which was one day after turning on the ZeBox powered air decontamination unit when the microbial load should have settled into a new level of equilibrium. The Shapiro-Wilk test (SW test) indicated that except for some data at position S1 (medicine table) and post deployment bacterial load at position S2/S3 (nurse station/ bed rails) in the single bed ICU, most data sets were not normally distributed (data available in supplementary material). In the multi bed ICU, only the post deployment surface bacteria at Position 1 and airborne bacterial load at position 3 and post deployment airborne bacterial load at position 4 were normally distributed. The results from the t test and the nonparametric “Mann-Whitney’s U test” (MWU test) provided similar levels of confidence. Results of the t-test for all the cases are shown in Tables S4 and S5, and that of the MWU test in tables S6 and S7. We see that the reduction brought about by deployment of ZeBox technology powered air decontamination unit is significant in nearly all the cases, except for surface bacteria at position 2 in the single bed ICU and surface fungi at position 4 in the multibed ICU. All details are available in the Supplementary material.

## Discussion

We carried out two independent studies to determine the efficiency of proprietary ZeBox technology powered air decontamination device in a single bed ICU and a multi bed ICU. Both studies were carried out when the rooms were occupied by patients, there was expected movement of hospital personnel and the hospital was fully functional. Devices underwent electrical safety and emission testing as per IEC60601-1-2 standards before deployment in clinical environments. Over the sampling interval, the baseline cultures in indoor air showed considerable variation with large standard deviations but less standard deviation in certain locations as evidenced through surface swab samples. Such variations have been previously described in the literature [32].

The robust and reproducible effect of the ZeBox technology powered air decontamination unit was observed on indoor air and frequently touched surfaces by microbial culture. After 3 hours of deployment of the unit, upwards of 95% reduction of bacterial load and upwards of 85% reduction of fungal load was observed in indoor air in both single bed ICU and multibed HICUs, so long as the sampling locations were within the effective range of the device. Similar extent of reduction was observed in surface bacterial and fungal loads. We thus showed that air decontamination could substantially and simultaneously reduce the levels of surface deposition in the same setting irrespective of the type of pathogen present, viz bacteria, fungi and their spores.

In the multibed HICU study, at sites distal to the deployed unit, the reduction in airborne bacterial load ranged from 24-45% and airborne fungal load from 35-70%. At the same distal sites, the surface bacterial load reduction ranged from 15-80% and fungal load reduction ranged from 61-73%. At sites proximal to the ZeBox technology powered air decontamination unit, the reduction of both bacterial and fungal load was greater than 95%. This indicates that a low bioburden zone was created with an approximate radius of 10-15 feet from the unit, for airborne bacterial and fungal load. These results are in concordance with the results obtained in earlier studies under controlled conditions [30].

To understand if the reduction in the microbial load post-deployment of ZeBox technology powered air decontamination unit was significant, we conducted a statistical analysis of the data (refer supplementary material for details). Since most statistical tests demand that the data be normally distributed, we first tested the data for normality using the Shapiro-Wilk test. We found that most of the datasets were *not* normally distributed. Therefore, to assess the significance of the reduction in microbial load due to ZeBox powered air decontamination unit, we conducted both a parametric test (Student’s t-test) which is applicable to normally distributed data, and a non-parametric test (Mann-Whitney’s U test) which is applicable to non-normally distributed data. Despite the different assumptions and theoretical basis underlying the two tests, their conclusions were the same. Except for two cases of surface microbes, one on the bed rail in the single-bed ICU and another at a location farthest from the ZeBox technology powered air decontamination unit in the multi-bed ICU, the tests yielded *p*-values significantly less than 0.05. This confirms that there was a significant reduction in the microbial load due to deployment of the ZeBox technology powered air decontamination unit.

Data on movement of people was not collected. Previously published studies linking occupancy of ICU to airborne culture numbers required intensive sampling over short time intervals which was not feasible in this study.

Several microbial strains resistant to Ceftazidine, Azithromycin and Ampicillin were found among the clinical isolates characterized. These are typical organisms found in hospital wards, some of which may be responsible for nosocomial or opportunistic infections in immunocompromised patients. The isolates were relatively more sensitive to the newer classes of antibiotics such as meropenem and linezolid. While most isolates were highly sensitive to Ciprofloxacin and Rifampicin, the latter is reserved as first line treatment for drug sensitive Tuberculosis, limiting its use against other infections. The *Kytococcus* and *Micrococcus* isolates which are resistant practically to all antibiotics tested except Linezolid would be expected to be particularly difficult to treat with available antibiotics, posing a challenge to infections in immunocompromised patients. Thus, an indoor environment equipped with an air decontamination unit which ideally eliminates microbes at source and provides near-sterile circulating air would be the desired way to prevent nosocomial infections.

Our study demonstrates that the innovative ZeBox technology can provide an effective trap and kill mechanism to eliminate a broad spectrum of airborne pathogens under clinical conditions. This in turn prevents re-settling of bacterial and fungal microorganisms on surfaces. Continuous operation of the ZeBox powered air decontamination unit can lead to ongoing reductions of pathogens in air and on environmental surfaces.

## Conclusion

Effective decontamination technology that aids infection control in healthcare spaces must do the following:

1. kill pathogenic or contaminating microbes instead of merely trapping

2. operate continuously and safely in human presence

3. and require near-zero manual intervention while operating close to the source of infection or contamination.

No other technology being evaluated globally meets all these requirements. While filtration technologies fail to meet the first criterion, UV and ionization-based technologies fail to meet the last two. The unique, extremely effective, energy-efficient technology, ZeBox satisfies all these attributes. The devices powered by the proprietary ZeBox technology effectively eliminate airborne microorganisms like *Staphylococcus aureus, Pseudomonas aeruginosa, Candida albicans, Aspergillus fumigatus spores, Mycobacterium smegmatis* [30] and *Mycobacterium tuberculosis*, bacteriophages such as MS2 phage and Phi X 174 (data not shown). Devices were previously shown to reduce 5log_10_ to 9log_10_ or 99.999-99.9999999% of viable microbial load based on the starting concentration under challenge conditions. In this study, we demonstrate that the ZeBox technology effectively eliminates the microbial population present in normally functioning hospital environments with efficiency over 95%, from the air and close to 85% from high contact surfaces like patient bed rails. Reducing the environmental microbial load will reduce the occurrence of nosocomial infections in healthcare environments. Although this study demonstrates the device’s capability in eliminating bacterial and fungal load from the environment, further study is required to assess impact on viruses under clinical settings, especially respiratory viruses. Nevertheless, this study successfully evaluates a novel decontamination technology that can be used not only in hospitals ICUs but also in other areas such as burns units and around immunocompromised patients, where the maintenance of low bioburden is critical to maintaining good health and preventing difficult to treat infections.

## Data Availability

The datasets supporting the conclusions of this article are included within the article.

## Abbreviations

|  |  |
| --- | --- |
| CDC | Centers for Disease Control and Prevention |
| CFU | Colony Forming Units |
| DNA | Deoxyribonucleic Acid |
| DMSO | Dimethyl sulfoxide |
| ESKAPE | Enterococcus faecium, Staphylococcus aureus, Klebsiella pneumoniae, Acinetobacter baumannii, Pseudomonas aeruginosa, and Enterobacter species |
| HAI | Hospital Acquired Infection |
| HBV | Hepatitis B Virus |
| HICU | High Intensity Care Unit |
| HSV | Herpes Simplex Virus |
| HVAC | Heating, Ventilation and Air Conditioning |
| ICU | Intensive Care Unit |
| MDR | Multi-Drug Resistant |
| MIC | Minimum Inhibitory Concentration |
| MRSA | Methicillin Resistant Staphylococcus Aureus |
| MWU test | Mann-Whitney’s U test |
| PBS | Phosphate-Buffered Saline |
| PCR | Polymerase Chain Reaction |
| rDNA | Recombinant DNA |
| rRNA | Ribosomal ribonucleic acid |
| SW test | Shapiro-Wilk test |
| UV | Ultra Violet |
| VRE | Vancomycin Resistant Enterococcus |

## Declarations

### A. Ethics approval and consent to participate

Institutional Ethics Committee approval were taken for this study. IEC Study Ref No 361/2017. for St John’s Hospital and Medical College, Bangalore, Karnataka, India and

### B. Consent for publication

All authors read and approved the final submitted version.

### C. Availability of data and material

The datasets supporting the conclusions of this article are included within the article.

### F. Authors’ contributions

SN, SS, BK, SC and IM designed the experiments, SJ, SB and DK performed the experiments, SP, DM and VN performed statistical analysis. VN and AG wrote the initial manuscript. AG managed funding.

## Acknowledgments

Authors acknowledges the kind help of staff members of Department of Microbiology, St John’s Hospital and Medical College, Bangalore, Karnataka, India and Department of Microbiology, Bangalore Baptist Hospital, Bangalore, Karnataka, India. We thank Dr. Santanu Datta and Dr.Janani Venkataraman for the constructive discussions on the manuscript.

## Supplementary material

**Supplementary Figure 1:**
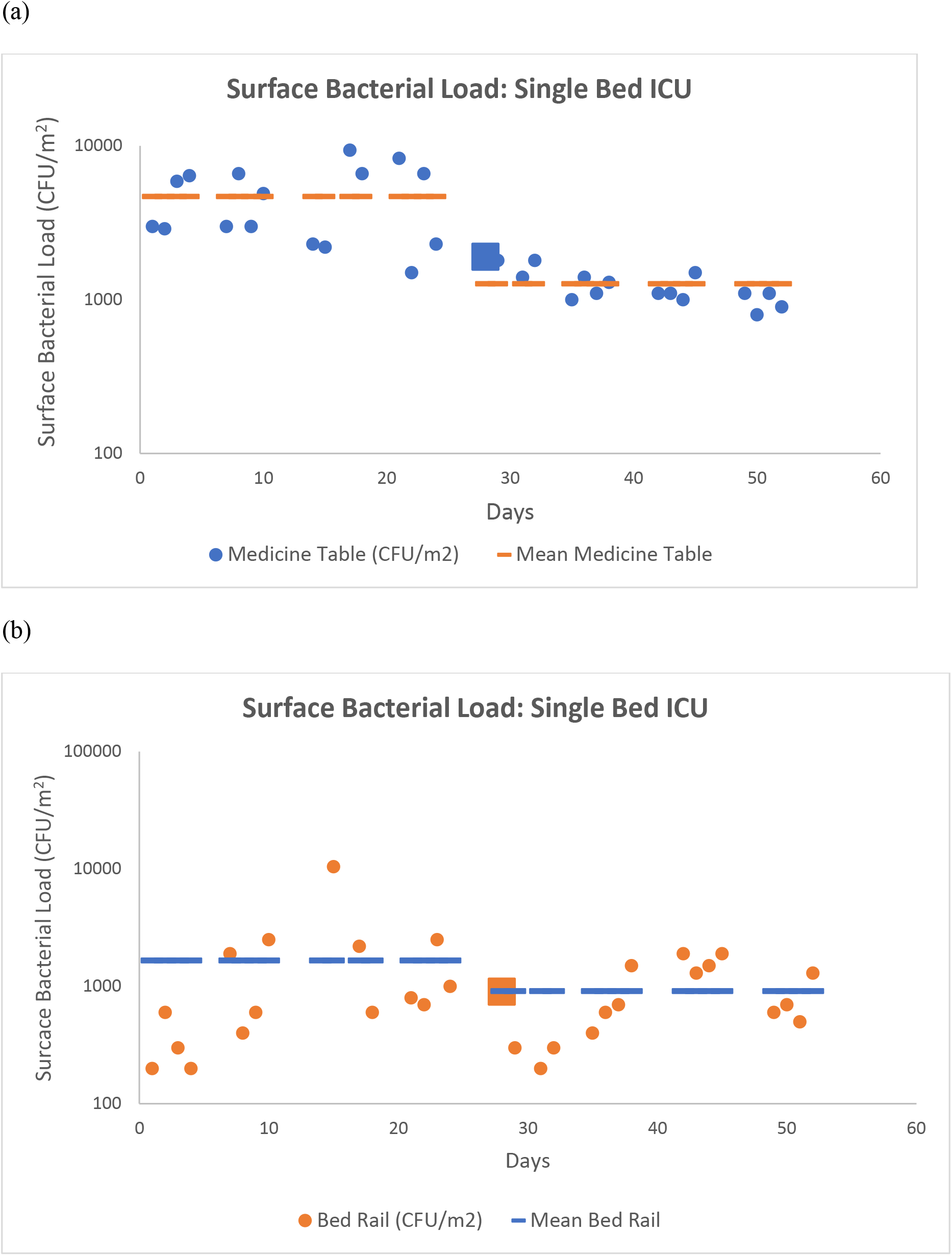
Surface Bacterial Loads in a Single bed ICU at two positions. (a) Position S1, Medicine table and (b) position S3, Bed rails. The average load before and after ZeBox deployment is depicted by a line for both positions. The ZeBox was deployed on day 28 and the first sample was taken within 3 hours of deployment. The microbial count for that time point is depicted by the square.

**Supplementary Figure 2:**
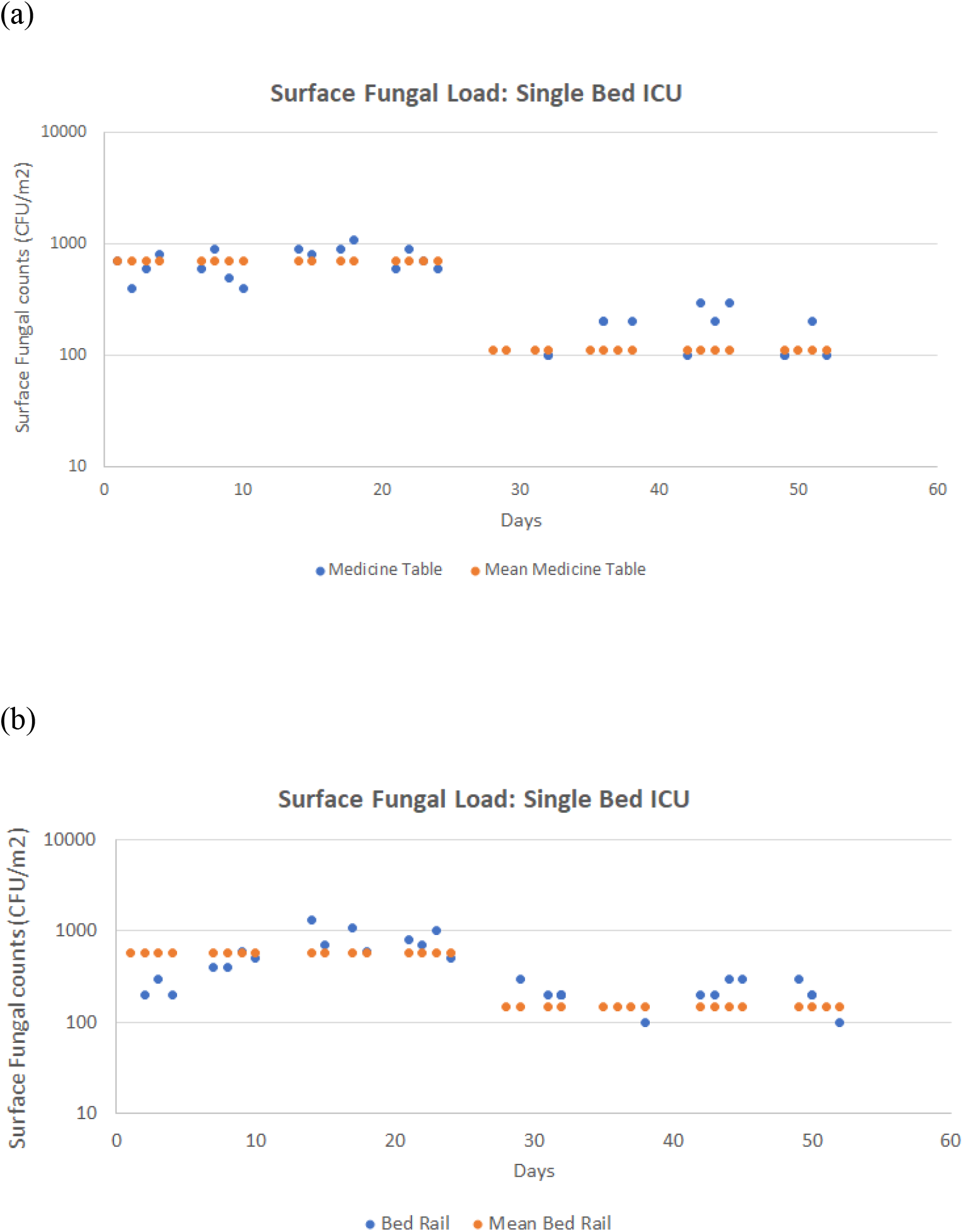
Surface Fungal Loads in a Single bed ICU at two positions. (a) Position S1, Medicine table and (b) position S3, Bed rails. The average load before and after ZeBox deployment is depicted by a line for both positions. The ZeBox was deployed on day 28 and the first sample was taken within 3 hours of deployment. The microbial count for that time point was zero at both positions and is not shown here.

**Supplementary Figure 3:**
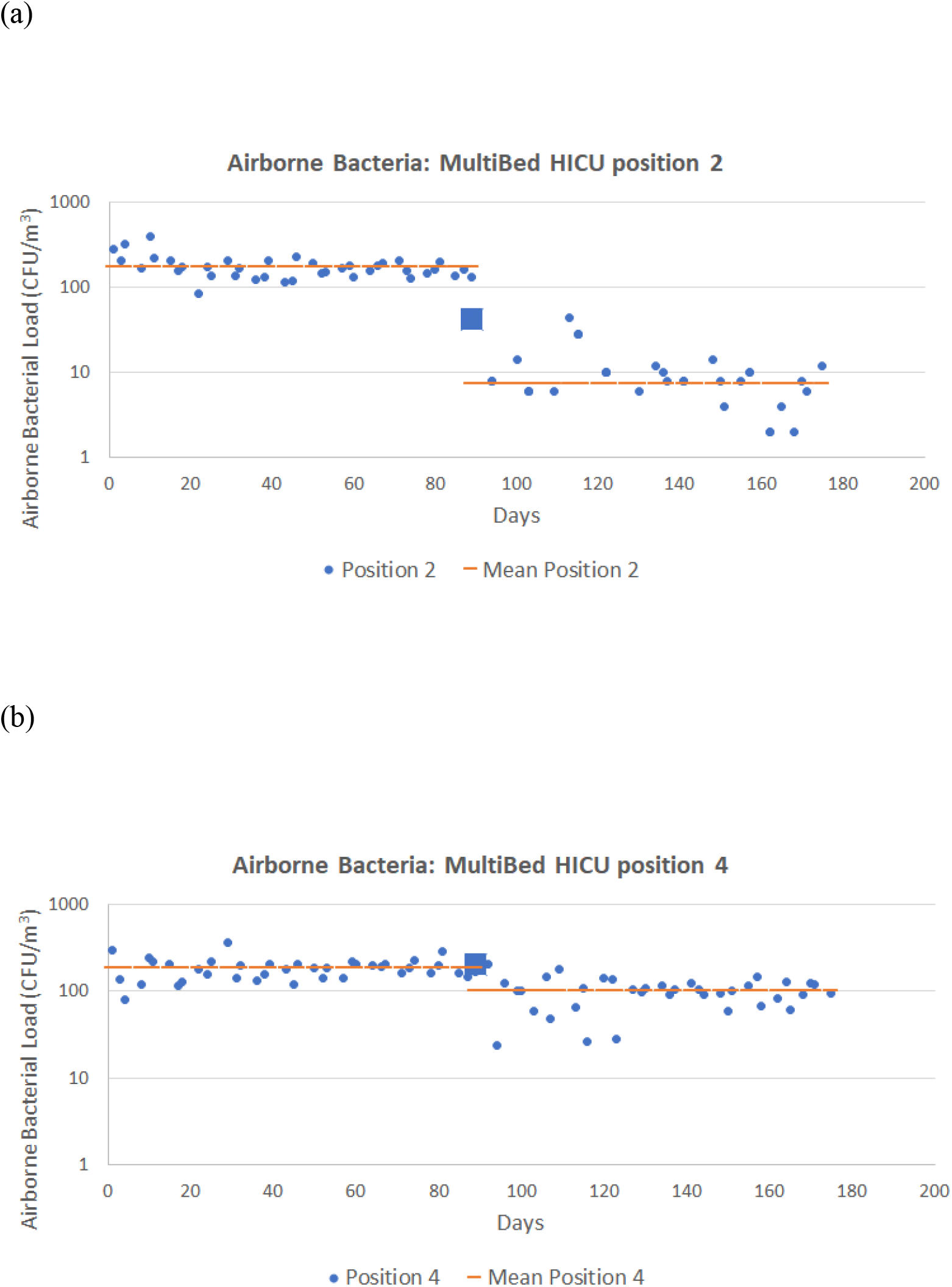
Airborne Bacterial load in Multi bed ICU. Active sampling was carried out from four positions. Positions 2 (a) was 8 feet away, and Position 4 (b) was 26 feet away from the ZeBox. The ZeBox was deployed on day 89 and the first sample was taken within 3 hours of deployment. The microbial count for that time point is depicted by a square.

**Supplementary Figure 4:**
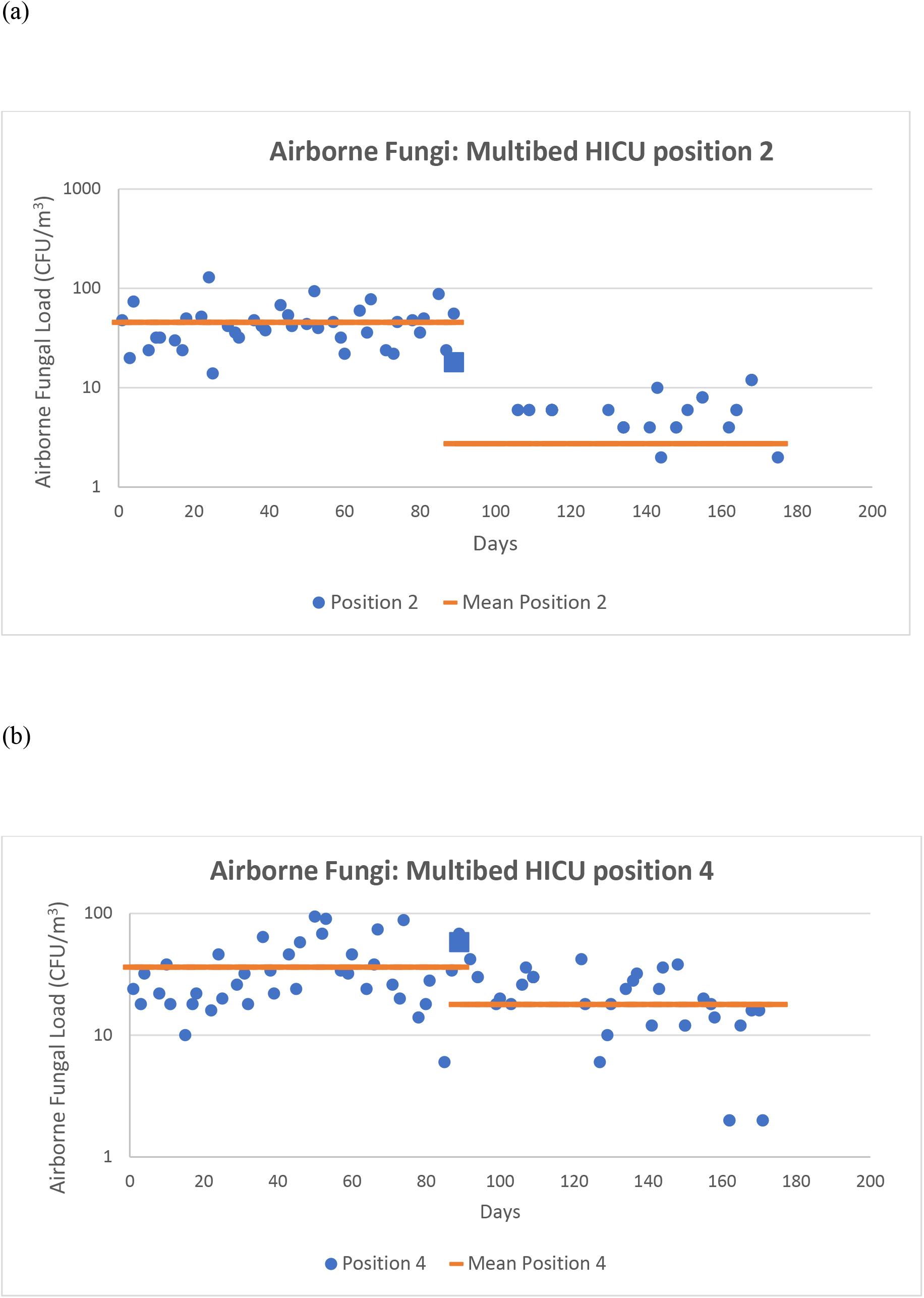
Airborne Fungal load in Multi bed ICU. Active sampling was carried out from four positions. Positions 2 (a) was 8 feet away, and Position 4(b) was 26 feet away from the ZeBox. The ZeBox was deployed on day 89 and the first sample was taken within 3 hours of deployment. The microbial count for that time point is depicted by a square.

**Supplementary Figure 5:**
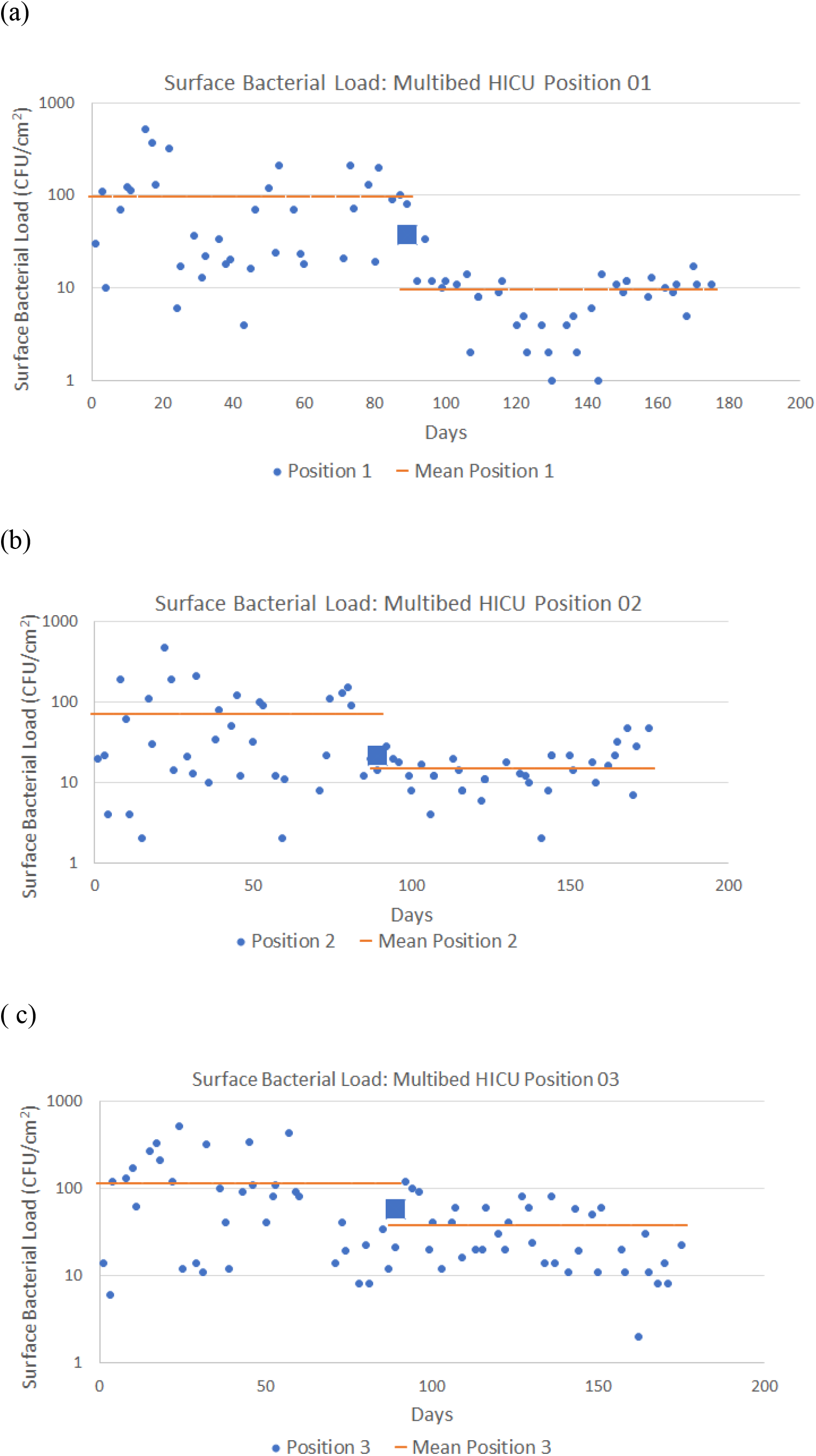
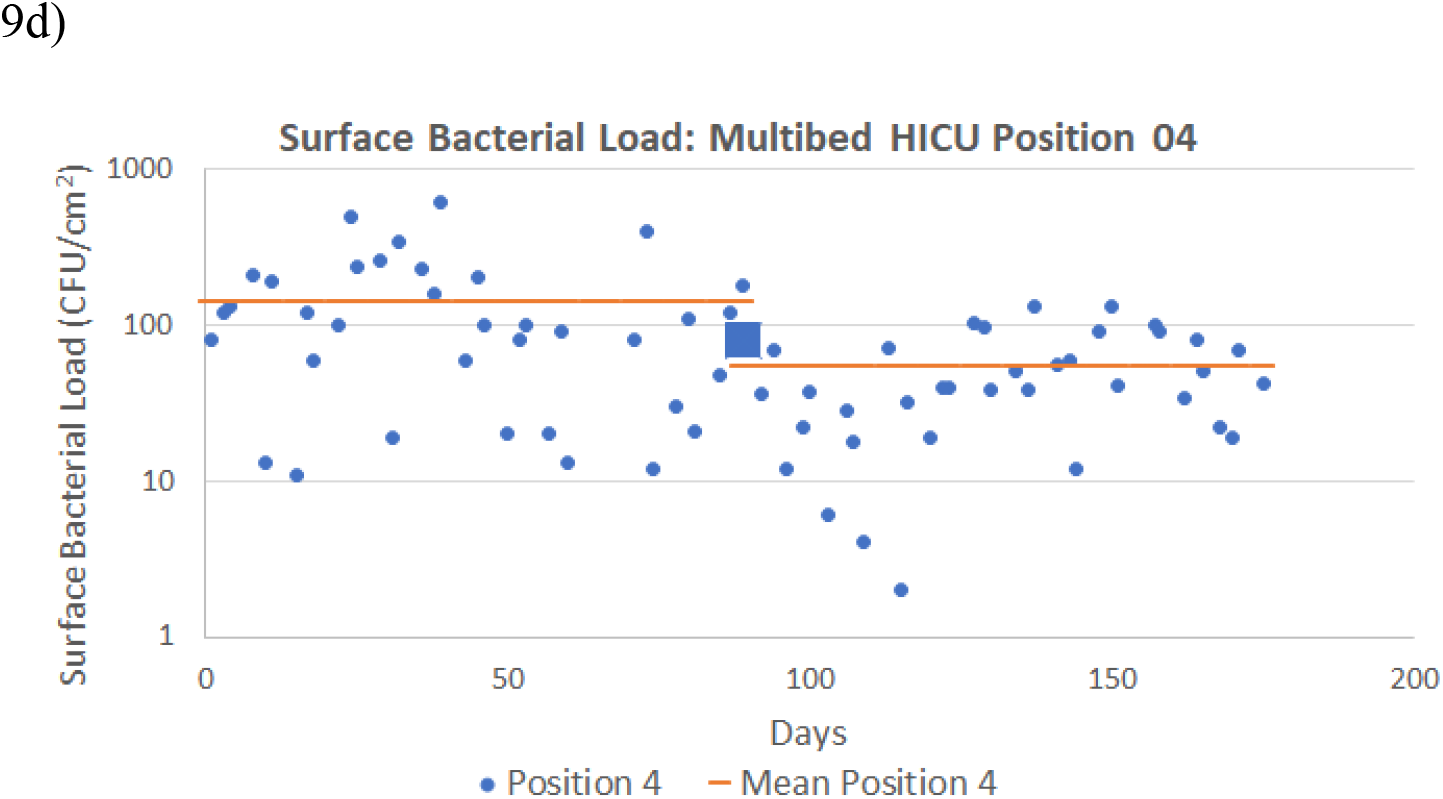
Surface Bacterial load in Multi bed ICU. Positions 1 and 2 were 2 feet and 8 feet away, respectively, from the ZeBox. Positions 3 (a) and 4 (b) were 25 feet and 26 feet away from the ZeBox. The ZeBox was deployed on day 89 and the first sample was taken within 3 hours of deployment. The microbial count for that time point is depicted by a square.

**Figure S4:**
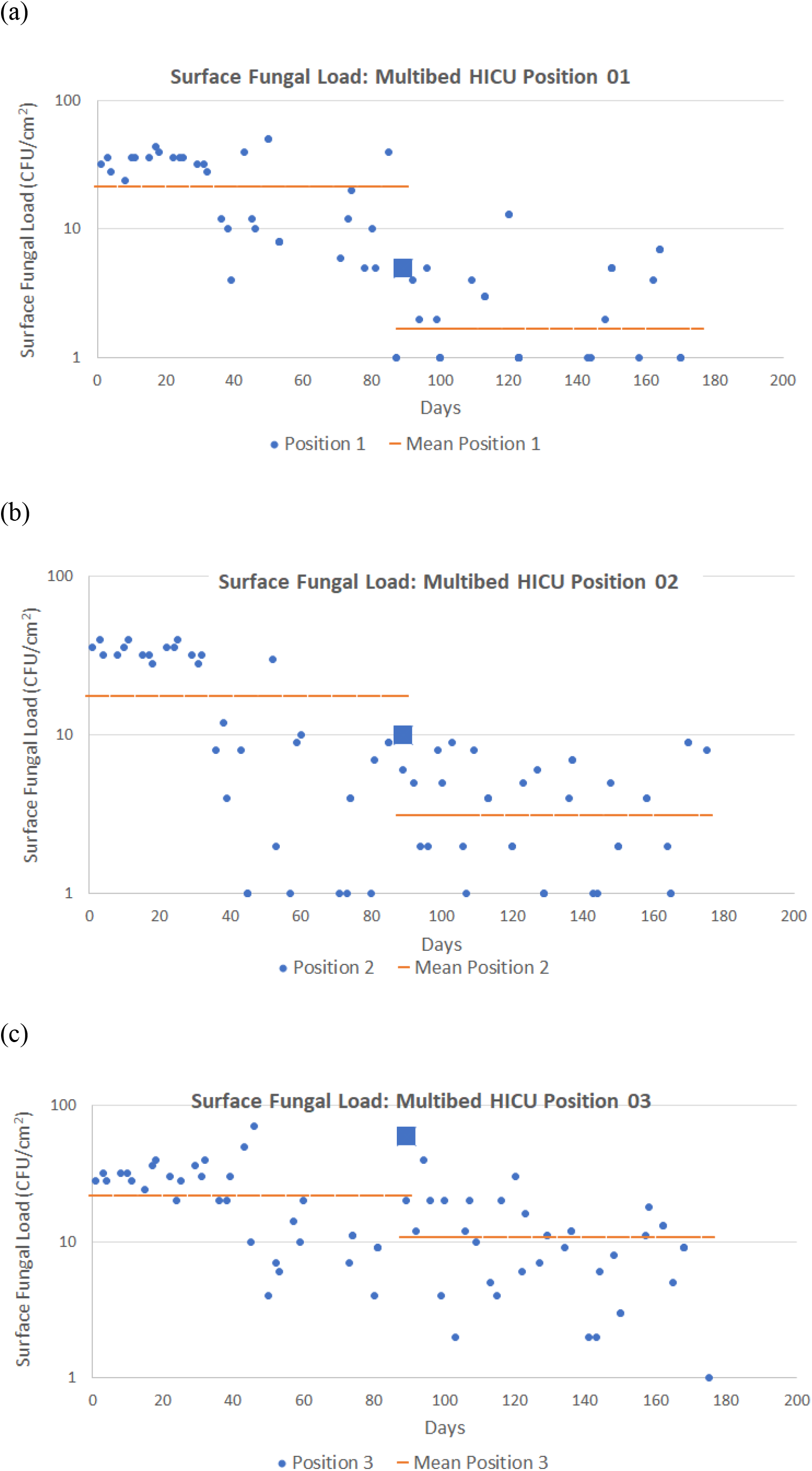
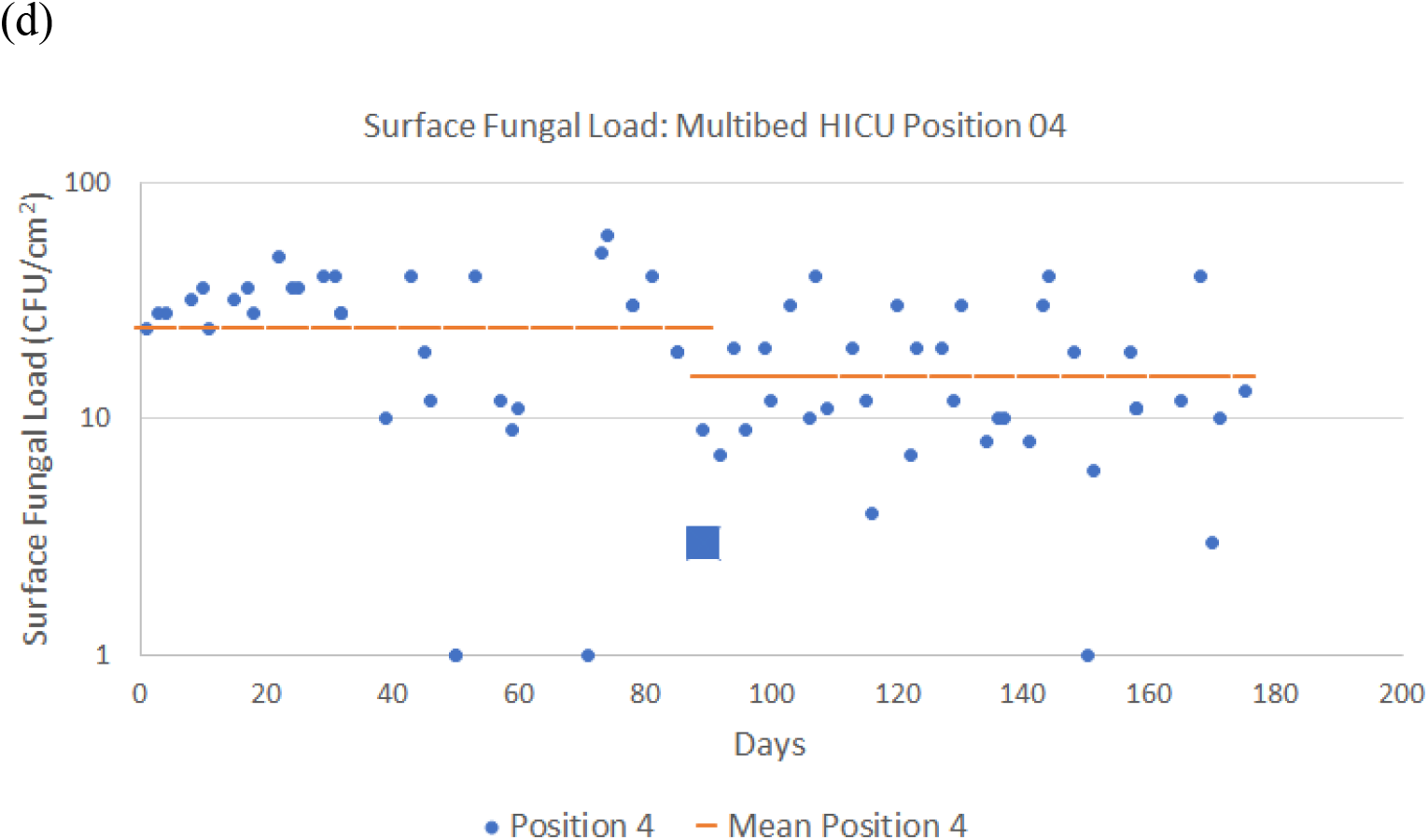
Surface Fungal load in Multi bed ICU. Positions 1(a) and 2 (b) were 2 feet and 8 feet away, respectively, from the ZeBox. Positions 3 (c) and 4 (d) were 25 feet and 26 feet away from the ZeBox. The ZeBox was deployed on day 89 and the first sample was taken within 3 hours of deployment. The microbial count for that time point is depicted by a square.

## Statistical analysis of data

Statistical tests were used to determine the significance of the reduction in microbial load due to ZeBox. We divide the time series of measured microbial load into pre-deployment and post-deployment periods. After turning on ZeBox, there is a finite period of transition until the microbial load settles to a new (lower) equilibrium level. The post-deployment period was reckoned to begin at the end of the transition period, assumed to be 1 day after turning on ZeBox.

Most statistical tests applicable to our case make two major assumptions: (1) The data in the pre- and post-deployment periods are statistically steady (or stationary); this implies that the statistics of the microbial load within a given period does not change progressively over time, and (2) The data is sampled from a normal (or, Gaussian) distribution. The first assumption is valid because we have left out the transition period from consideration. Whether the second assumption is valid was determined by testing the data for normality using Shapiro-Wilk test (SW test). The null hypothesis for the SW test is that the dataset is sampled from a normal distribution. Table S1 and Table S2 show the results of the SW test; each entry is a pair of the SW-statistic and the corresponding p-value. All the statistical tests were done using the **scipy-**statistics package in **python**. The cases with p-value > 0.05, marked in green, indicate that the corresponding datasets are indeed sampled from a normal distribution. We see that most datasets are in fact not sampled from a normal distribution.

**Table S1.**
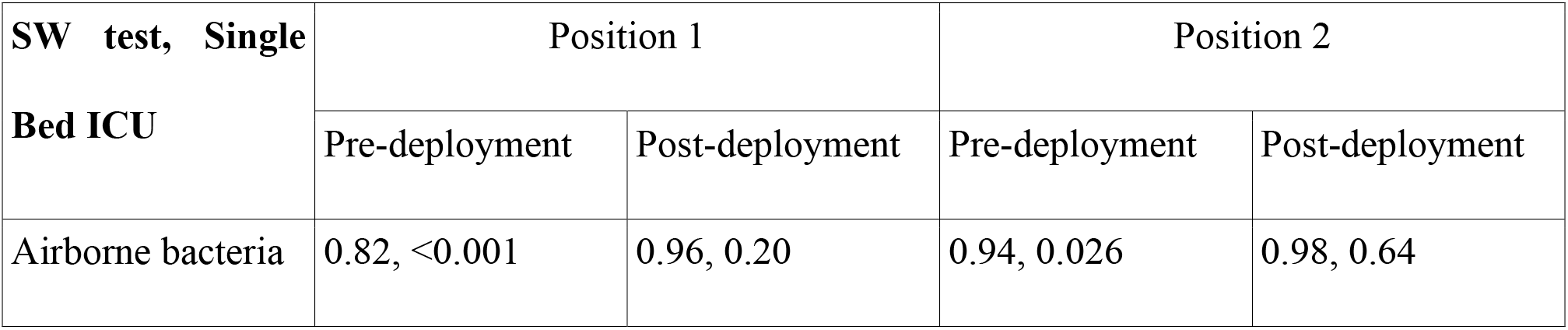
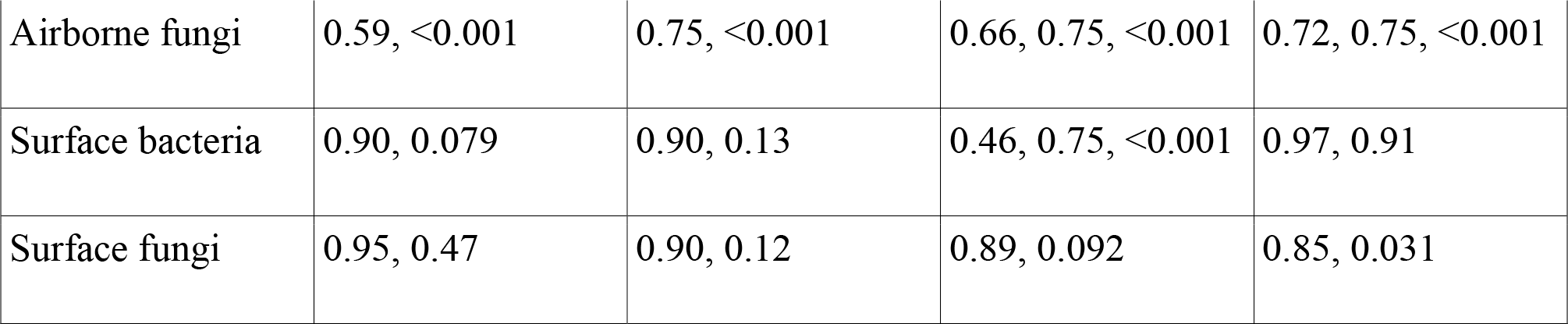
SW test for single bed ICU data. Each entry is a pair of SW-statistic and the corresponding p-value.

| SW test, Single<br>Bed ICU | Position 1 |  | Position 2 |  |
| --- | --- | --- | --- | --- |
|  | Pre-deployment | Post-deployment | Pre-deployment | Post-deployment |
| Airborne bacteria | 0.82, <0.001 | 0.96, 0.20 | 0.94, 0.026 | 0.98, 0.64 |
| Airborne fungi | 0.59, <0.001 | 0.75, <0.001 | 0.66, 0.75, <0.001 | 0.72, 0.75, <0.001 |
| Surface bacteria | 0.90, 0.079 | 0.90, 0.13 | 0.46, 0.75, <0.001 | 0.97, 0.91 |
| Surface fungi | 0.95, 0.47 | 0.90, 0.12 | 0.89, 0.092 | 0.85, 0.031 |

**Table S2.**
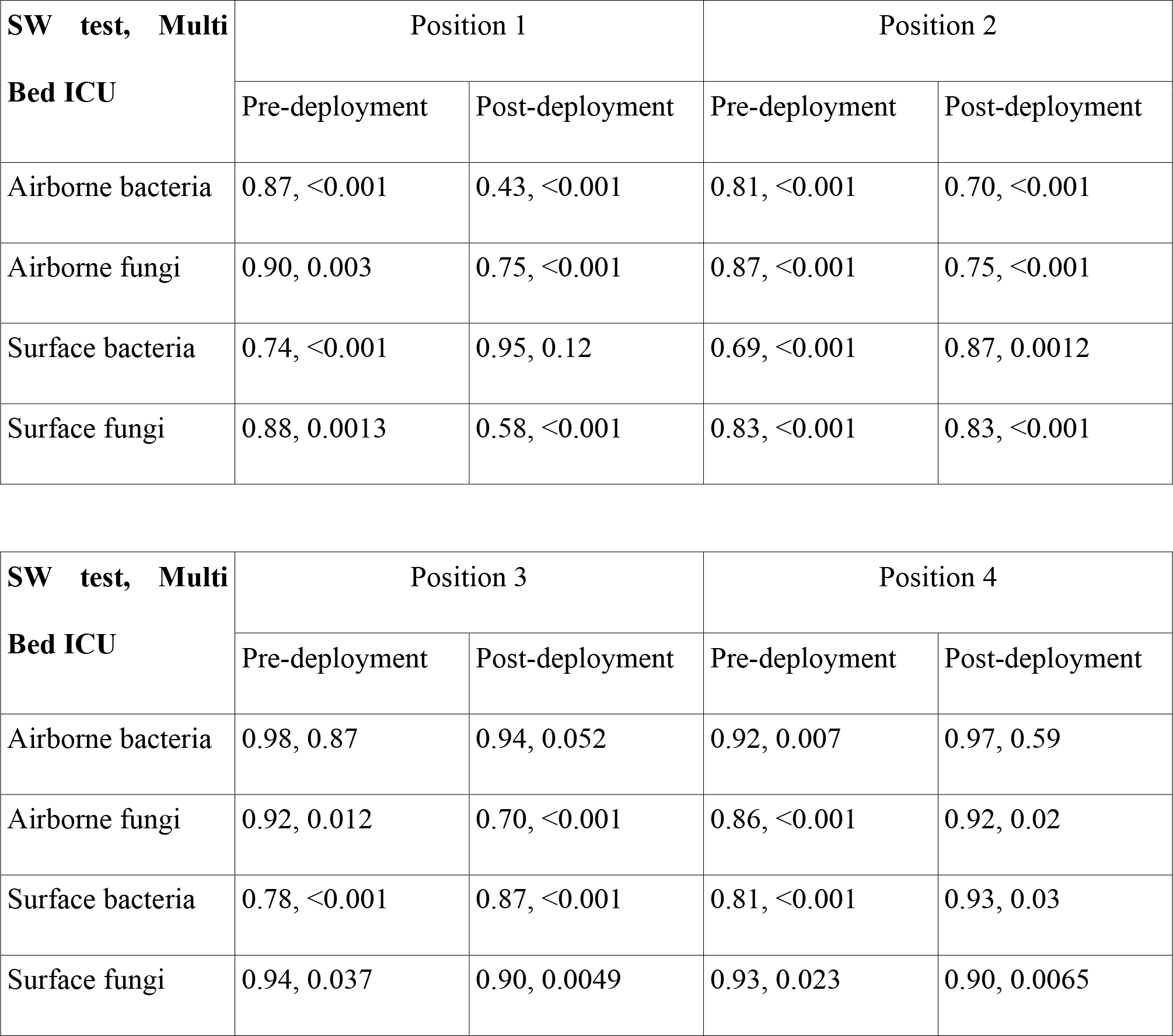
SW test for multi bed data. Each entry is a pair of SW-statistic and the corresponding p-value.

If pre- and post-deployment datasets are both normally distributed then, to assess the significance of the reduction in microbial load post-deployment of ZeBox, we may use a parametric test such as the “two sample, left tailed t-test”, otherwise a non-parametric test such as the “Mann-Whitney’s U test” (MWU test) is appropriate [1]. For the t-test, the null hypothesis is that the mean microbial load in pre- and post-deployment periods are the same, and the alternative hypothesis is that the ZeBox brings about a reduction in the mean microbial load (which therefore requires a left-sided test). For the MWU test, the null hypothesis is that the pre- and post-deployment datasets are sampled from the same probability distribution, while the alternative hypothesis is that they are sampled from different distributions and that the ZeBox brings about a reduction in the microbial load (which therefore requires a left-sided test).

Results of the t-test for all the cases are shown in tables S3 and S4, and that of the MWU test in tables S5 and S6. We see that the reduction brought about by deployment of ZeBox is significant in nearly all the cases, except for surface bacteria at position 2 in the single bed ICU and surface fungi at position 4 in the multi bed ICU

**Table S3.** Two sample, left-sided t-test for single bed data. Each entry is a pair of t-statistic and the corresponding p-value.

| **t-test, Single Bed ICU** | Position 1 | Position 2 |
| --- | --- | --- |
| Airborne bacteria | 14.4, <0.001 | 14.4, <0.001 |
| Airborne fungi | 6.8, <0.001 | 6.9, <0.001 |
| Surface bacteria | 5.1, <0.001 | 1.2, 0.13 |
| Surface fungi | 9.3, <0.001 | 4.3, <0.001 |

**Table S4.**
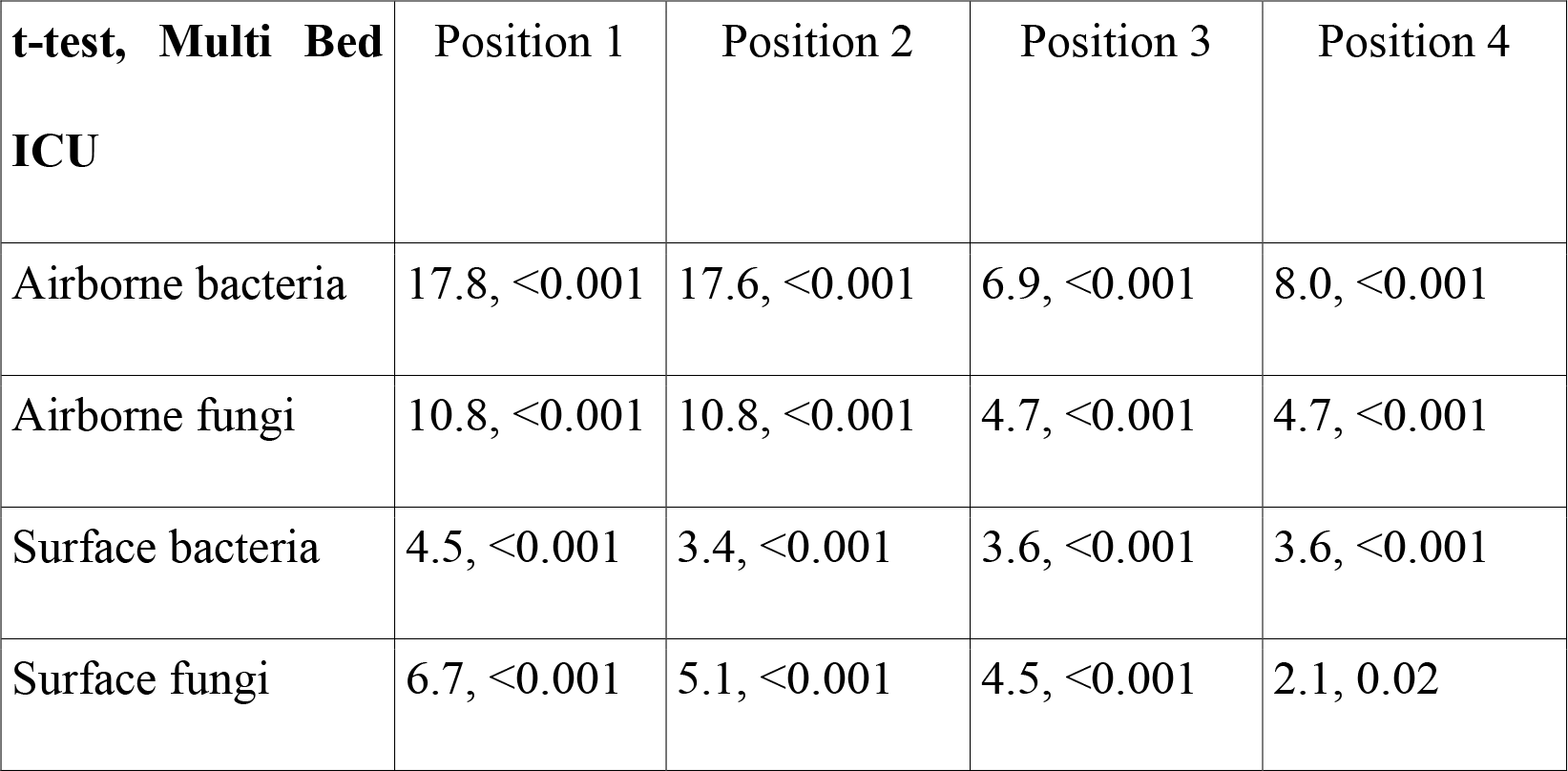
Two sample, left-sided t-test for multi bed data. Each entry is a pair of t-statistic and the corresponding p-value.

**Table S5.** Two sample, left-sided MWU-test for single bed data. Each entry is a pair of MWU-statistic and the corresponding p-value.

| **MWU-test, Single Bed ICU** | Position 1 | Position 2 |
| --- | --- | --- |
| Airborne bacteria | 1591, <0.001 | 1570, <0.001 |
| Airborne fungi | 1580, <0.001 | 1541, <0.001 |
| Surface bacteria | 207, <0.001 | 111, 0.39 |
| Surface fungi | 208, <0.001 | 186, <0.001 |

**Table S6.**
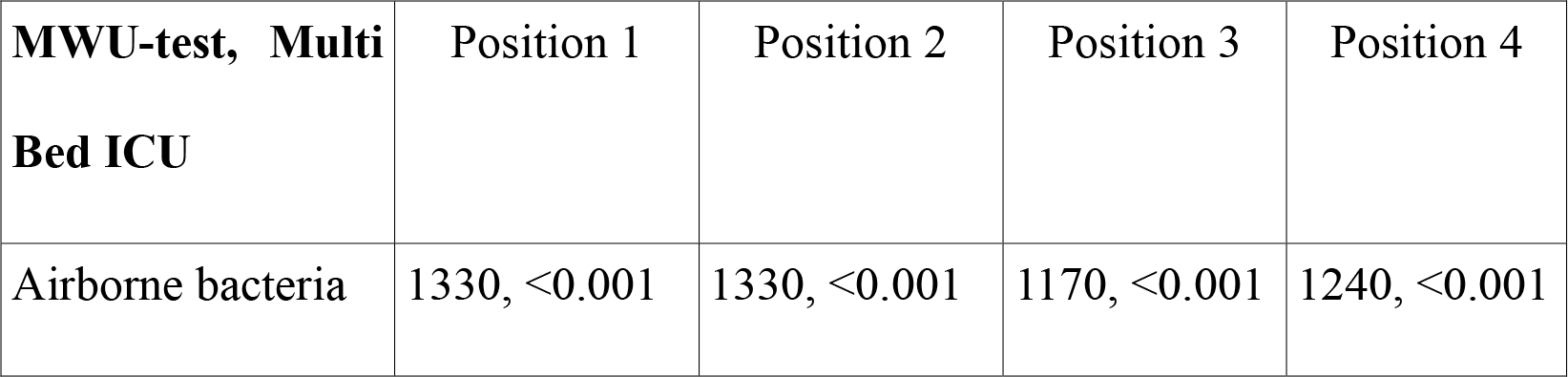
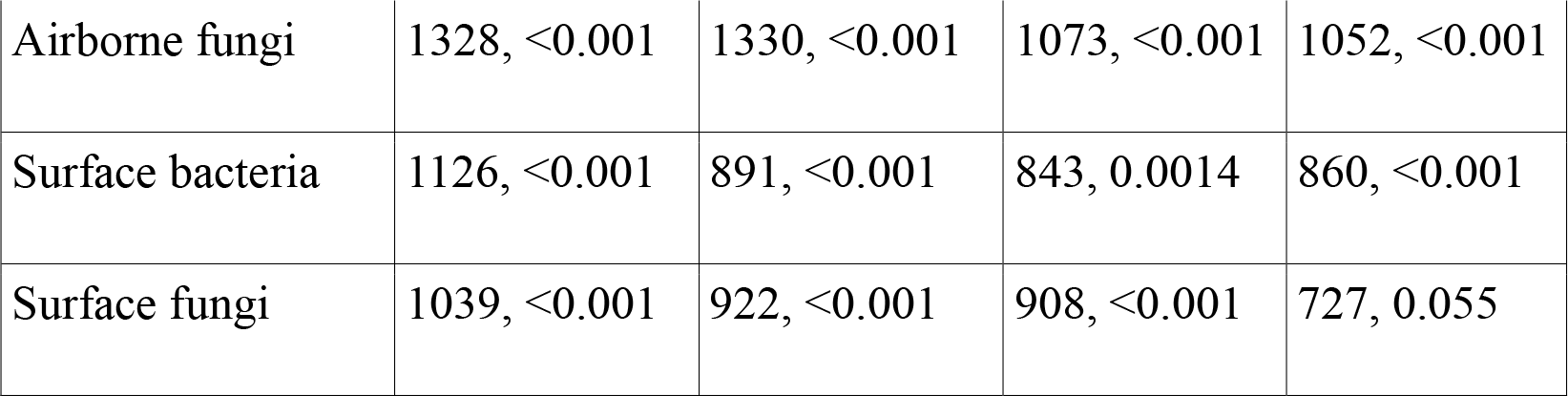
Two sample, left-sided MWU-test for multi bed data. Each entry is a pair of MWU-statistic and the corresponding p-value.

| **MWU-test, Multi Bed ICU** | Position 1 | Position 2 | Position 3 | Position 4 |
| --- | --- | --- | --- | --- |
| Airborne bacteria | 1330, <0.001 | 1330, <0.001 | 1170, <0.001 | 1240, <0.001 |
| Airborne fungi | 1328, <0.001 | 1330, <0.001 | 1073, <0.001 | 1052, <0.001 |
| Surface bacteria | 1126, <0.001 | 891, <0.001 | 843, 0.0014 | 860, <0.001 |
| Surface fungi | 1039, <0.001 | 922, <0.001 | 908, <0.001 | 727, 0.055 |

